# Prediction of response to transcranial magnetic stimulation treatment for depression using electroencephalography and statistical learning methods, including an out-of-sample validation

**DOI:** 10.1101/2023.10.24.23297492

**Authors:** Neil W Bailey, Ben D. Fulcher, Martijn Arns, Paul B Fitzgerald, Bernadette Fitzgibbon, Hanneke van Dijk

## Abstract

**Background:** Repetitive transcranial magnetic stimulation (rTMS) has shown efficacy for treating depression, but not for all patients. Accurate treatment response prediction could lower treatment burden. Research suggests machine learning trained with electroencephalographic (EEG) data may predict response, but only a limited range of measures have been tested.

**Objectives:** We used >7000 time-series features to comprehensively test whether rTMS treatment response could be predicted in a discovery dataset and an independent dataset.

**Methods:** Baseline EEG from 188 patients with depression treated with rTMS (125 responders) were decomposed into the top five principal components (PCs). The *hctsa* toolbox was used to extract 7304 time-series features from each participant and PC. A classification algorithm was trained to predict responders from the feature matrix separately for each PC. The classifier was applied to an independent dataset (*N* = 58) to test generalizability on an unseen sample.

**Results:** Within the discovery dataset, the third PC (which showed a posterior-maximum and prominent alpha power) showed above-chance classification accuracy (68%, *p*_FDR_ = 0.005, normalised positive predictive value = 114%). Other PCs did not outperform chance. The model generalized to the independent dataset with above-chance balanced accuracy (60%, *p* = 0.046, normalised positive predictive value = 114%). Analysis of feature-clusters suggested responders showed more high frequency power relative to total power, and a more negative skew in the distribution of their time-series values.

**Conclusion:** The dynamical properties of PC3 predicted treatment response with moderate accuracy, which generalized to an independent dataset. Results suggest treatment stratification from pre-treatment EEG may be possible, potentially enabling better outcomes than ‘one-size-fits-all’ treatment approaches.

## Introduction

Repetitive transcranial magnetic stimulation (rTMS) has become an increasingly common treatment for treatment-resistant depression. However, meta-analyses suggest response rates to rTMS are <50% (Berlim et al., 2014; Cao et al., 2018; Sehatzadeh et al., 2019). Clinical observation data suggests that patients who respond to rTMS often show large improvements, while non-responders often show minimal change in depression severity, so that response patterns are of a bimodal distribution (Fitzgerald et al., 2016). This pattern provides motivation to investigate whether there is a way to predict treatment responses, and thus reduce the burdens to patients and clinical costs when patients would fall into the category of ‘non-responder’ (Kar, 2019). Achieving this goal could be a first step towards ‘stratified psychiatry’ for depression, where multiple predictive measures of response to a range of different treatments could guide treatment selection and lead to more successful depression treatment responses as patients are provided with the treatment they are most likely to respond to (Arns et al., 2022).

Since depression is associated with underlying brain function (de Aguiar Neto & Rosa, 2019; Drevets et al., 2008), it is sensible to assess neural activity as a potential predictor of treatment response, which is commonly measured using functional magnetic resonance (fMRI) (Ge et al., 2020). However, although baseline fMRI measures have sometimes been highly accurate at predicting responders (Cash et al., 2019; Drysdale et al., 2017), replication attempts have suggested that prediction accuracy may have been due to data-processing artifacts, and replication attempts have not been successful (Dinga et al., 2019; Hopman et al., 2021). fMRI is also expensive, and as such fMRI measures may be clinically impractical for response prediction (Watts et al., 2022). In contrast, EEG activity provides an attractive alternative as a potential predictive measure. EEG research has shown promise for detecting the differences in neural activity associated with depression (de Aguiar Neto & Rosa, 2019), and is cost-effective, practical, feasible, and scalable to implement in a clinical setting (Watts et al., 2022; Widge et al., 2019). The feasibility and practicality of recording EEG in a clinical setting has been demonstrated by the TDBRAIN dataset, where >1400 EEG recordings from a clinical practice have been made available, including EEG recordings obtained prior to rTMS treatment for depression (van Dijk, van Wingen, et al., 2022). In addition to its feasibility, EEG is recorded from the outer layer of the cortex where rTMS treatment is applied, and perhaps for this reason, meta-analysis has indicated rTMS response prediction to be more accurate than prediction of antidepressant response to other treatments (Watts et al., 2022).

Furthermore, there is preliminary evidence that prospective response predictions obtained from baseline EEG data can increase the efficacy of antidepressant treatment recommendations in a clinical setting (in an open label study; van der Vinne et al. (2021)). With the availability of large datasets like TDBRAIN, statistical classification models including machine learning hold promise for accurately predicting treatment response. Machine learning is an umbrella term for the set of techniques and algorithms that automatically learn patterns from data, without explicit programming for each application (Watts et. al., 2022). The classification subset of machine learning approaches then use the learned model to predict the label of unseen data (Watts et al., 2022). Meta-analysis of machine learning studies of the prediction of response to depression treatments has suggested an overall mean accuracy of 83.93% and an average sensitivity and specificity of 78% and 85% respectively, most commonly from single datasets without using a validation dataset, posing the risk that prediction accuracies are inflated by improperly handled data leakage between the algorithm training set and the accuracy test set (Watts et al., 2022).

Despite these optimistic results, prediction research has not yet been applicable in clinical practice. This is likely because studies still suffer from small sample sizes and a lack of cross-validation and so positive results may simply reflect overfitting, and may not generalise well (Watts et al., 2022; Widge et al., 2019). Indeed, very few independent replications have been reported. When independent replications are reported, only a few have been successful (Corlier, Carpenter, et al., 2019; Roelofs et al., 2021). Replication attempts have often been unsuccessful (Widge et al., 2013), including our own research, (Bailey et al., 2021; Krepel et al., 2018). One potential reason for non-replication is that the vast majority of the original studies have not used an independent dataset to verify the generalizability of their prediction model and, as such, the studies cannot rule out that the possibility that their results are due to the model over-fitting to characteristics specific to the dataset they were trained on (Arns et al., 2016; Squarcina et al., 2021; van der Vinne et al., 2021). That is, the prediction accuracy may depend on patterns in the discovery dataset, which may not generalise to another dataset, thus not providing the potential for clinical applicability (van Dijk, Koppenberg, et al., 2022; Watts et al., 2022). The lack of successful replications highlights the vital importance of verifying the generalizability of trained prediction models on independent samples (van Dijk, Koppenberg, et al., 2022). Prediction studies that have used a within-study replication or independent validation datasets have shown more modest (but more trustworthy) predictive accuracy (Krepel et al., 2020).

Additionally, while some of the most successful predictions have been produced by complex models with many parameters (such as deep-learning algorithms), these approaches have limitations, and these complex models are not always superior for providing accurate predictions (Mignan & Broccardo, 2019). In particular, the aspects of the data and inner workings of the algorithms that provide accurate predictions are often relatively opaque, and as such, difficult to implement and reproduce (Traut et al., 2022; Van Der Donckt et al., 2022). Due to the large number of parameters that can be tuned, the risk of over-fitting is also high (Traut et al., 2022). Further, the opaque nature of the deep-learning approach means it is often difficult to interpret which aspects of the data lead to accurate predictions, leaving researchers and clinicians unable to understand how the algorithm discerns responders and non-responders (Squarcina et al., 2021; Van Der Donckt et al., 2022). Given these limitations, a recent review of the literature identified only four potentially robust biomarkers for the prediction of response to non-invasive brain stimulation (two fMRI biomarkers and two EEG biomarkers) (Klooster et al., 2023). However, given the heterogeneity across individuals with depression, and the likely subtle relationship between any baseline marker and treatment response, it may be that single biomarkers will be unable to obtain the predictive accuracy required for clinical implementation. As such, it has been suggested that a multivariate approach, which takes into account many different characteristics of the data, might be necessary. As such, there is a “critical need for a systematic comparison of several types of features” (Watts et al., 2022).

To address these issues, we used the highly comparative time-series analysis (*hctsa*) software to measure >7000 statistical properties (or features) of univariate EEG time-series data from responders and non-responders to rTMS treatment for depression (Fulcher & Jones, 2017; Fulcher et al., 2013). We then trained a machine learning algorithm on these features to obtain a model that enabled out-of-sample predictions of responder and non-responders to rTMS treatment. Finally, we performed an individual feature analysis to provide an indication of both which analysis methods best separated responders and non-responders (Fulcher & Jones, 2017; Fulcher et al., 2013). In contrast to the relative opaqueness of deep-learning approaches, the time-series features analysed in the current study are the result of transparent algorithms grounded in interpretable time-series theory that can thus provide understanding of the types of time-series structures that best distinguish the EEG dynamics of responders and non-responders. Within EEG research, *hctsa* has previously been used to provide a data-driven sleep stage categorization from EEG data and to detect the higher order features that separate those sleep stages, allowing researchers to gain new insights into how sleep stages differ (Decat et al., 2022). It has also been used to detect seizure activity (Fulcher et al., 2013), and to provide novel understanding of how brain activity differs in long-term meditators (Bailey et al., 2023), as well as being applied in other fields such as in fMRI research, astrophysics, and in energy monitoring. The toolbox includes features derived from the Fourier power-spectrum, and other linear correlation-based statistics that are commonly used in EEG research, but also includes a much broader range of methods that quantify entropy, fractal scaling, stationarity, and many other types of features (Fulcher & Jones, 2017; Fulcher et al., 2013). *hctsa* computes features from a single time-series from each participant as its input, and EEG data is typically recorded as many time-series (from many electrodes). As such, we used a principal component analysis (PCA) dimensionality-reduction technique to extract a smaller number of components explaining a maximum amount of variance from time-series for analysis, while preserving the majority of the variance in the full multivariate time-series dataset (Bailey et al., 2023).

Given the aforementioned background, our aim was to use *hctsa* to determine whether a simple feature-based time-series classification model applied to a comprehensive feature set from EEG time-series data (without any hyperparameter tuning) could accurately predict response to rTMS treatment for depression from pre-treatment data. We aimed to test the approach using leave-one-participant-out cross-validation in a discovery dataset, and verify its generalizability to an unseen, independent validation dataset. For the independent validation dataset test we implemented a recommended “blinded” approach, whereby the primary author (NWB) submitted the prediction response categories for the validation dataset to other authors (MA and HvD) for them to calculate the prediction accuracy results, such that NWB is still not aware of the response categories of the out-of-sample dataset (Arns et al., 2022). Based on the conservative view of previous research, we hypothesized that we would obtain modest prediction accuracy. Additionally, we aimed to use the interpretability of the *hctsa* approach to characterise the best predictive features from the baseline EEG data. Since no previous research has used such a comprehensive list of time-series analyses as is provided by *hctsa*, we hypothesized that the analysis would provide us with a novel list of the features that could be predictive of rTMS response, and that this list might contain features that have not previously been used in EEG analysis e.g., that may be more commonly used in fields like seismology or economics.

## Methods

### Data

We used a subset of the full publicly available TDBRAIN dataset that contained participants over 18 years of age (*N* = 968), to optimize pre-processing parameters for the application of *hctsa* to EEG data (referred to as the ‘parameter-testing’ dataset, with the full explanation of this is reported in the supplementary materials). Then, for our primary analysis, we used a subset of the TDBRAIN dataset which included 196 participants who had been treated with rTMS to the DLPFC (98 male, 18 to 78 years of age, M = 43.62, SD = 12.82). Participants within this dataset had a diagnosis of non-psychotic major depressive disorder or dysthymia and a Beck Depression Inventory (BDI-II-NL) > 14 at baseline. Participants were treated with either 10Hz left hemisphere rTMS (*N* = 74), 1Hz right hemisphere rTMS (*N* = 115), or bilateral rTMS, applied as both 10Hz left and 1Hz right hemisphere rTMS (*N* = 7) (Donse et al., 2018, Van Dijk et al. 2022). Participants received treatment with at least 10 sessions of rTMS over the DLPFC or until response within these 10 sessions. Exclusion criteria for the rTMS sample were prior ECT treatment, epilepsy, traumatic brain injury, a current psychotic disorder, wearing a cardiac pacemaker, metal parts in the head, or pregnancy. Mean BDI-II-NL score at baseline was M = 31.26 (SD = 10.02). EEG recordings were obtained with participants seated in a sound and light attenuated room at an ambient temperature of 22°C. EEG data were acquired from 26 electrodes: Fp1, Fp2, F7, F3, Fz, F4, F8, FC3, FCz, FC4, T3, C3, Cz, C4, T4, CP3, CPz, CP4, T5, P3, Pz, P4, T6, O1, Oz and O2 (using either Quikcap or ANT WaveGuard caps; NuAmps; 10-20 electrode international system). EOG and ECG electrodes were also applied, but for the current study these were removed from the data prior to pre-processing. Two minutes of EEG data was recorded while participants rested with their eyes closed (EC). The operator did not intervene when drowsiness patterns were observed in the EEG. Data were referenced to averaged mastoids with a ground at AFz. Skin resistance was <10kΩ for all electrodes, and data were recorded at a sampling rate of 500Hz, and low-pass filtered at 100 Hz prior to digitization. After exclusion of eight participants during pre-processing (explained below), 188 participants were present in the data, 125 of whom were classified as responders, defined as a 50% reduction in BDI-II-NL scores from the baseline timepoint until the end of treatment. After exclusions, the final sample of participants included those treated with 10Hz left hemisphere rTMS (*N* = 71), 1Hz right hemisphere rTMS (*N* = 110), or bilateral 10Hz left and 1Hz right hemisphere rTMS (*N* = 7). All participants received the EEG as part of their routine care and provided informed consent for their data to be recorded and shared for the purposes of research.

### EEG pre-processing

To estimate suitable EEG pre-processing steps prior to submitting the rTMS data to the *hctsa*, we tested how accurately we could predict the sex of participants of using a larger parameter-testing dataset (N = 968, also obtained from TDBRAIN) by varying the pre-processing parameters and performing a manual search of sex prediction accuracy outcomes as the target to be optimized. This helped us determine effective artifact cleaning, epoch length, EEG sampling rate, PCA decomposition and PCA component inclusion parameters for predictions based on EEG data prior to implementing *hctsa* for prediction of response to rTMS. We have briefly reported the options and results of these parameter tests in our supplementary materials.

To pre-process the EEG data, we used the automatic EEG cleaning toolbox RELAX to clean the data (Bailey et al., 2022a; Bailey et al., 2022b). First, data was high-pass filtered using a fourth-order Butterworth filter at 1Hz, and a notch filter was applied from 47 to 53Hz. PREP’s automatic bad electrode detection and removal method was used (Bigdely-Shamlo et al., 2015), followed by RELAX’s default bad electrode detection and removal approach. Extreme outlying data periods were marked for exclusion from the Multi-channel Winer Filtering (MWF) (Somers et al., 2018) and were rejected from the data prior to independent component analysis (ICA) using RELAX’s default method. Two rounds of MWF were applied using a delay period of 12 (so that 24 samples or 48ms surrounding each timepoint were taken into account when constructing the artifact and clean data templates and when applying the spatial-temporal MWF cleaning to the data), first cleaning muscle activity, then horizontal eye movements and drift together (since data were recorded while participants had their eyes closed, no blink cleaning was required) (Somers et al., 2018). Low-pass filtering was then applied to the data at 80Hz, and robust average re-referencing was applied to the data (as independent component analysis performs better under these conditions). Fast Independent Component Analysis (ICA) was performed on the data to separate the data into its underlying independent components (Hyvarinen, 1999). Wavelet enhanced ICA (Castellanos & Makarov, 2006) was used to reduce artifactual components identified by ICLabel, before data were reconstructed into the scalp space (Pion-Tonachini et al., 2019).

After this pre-processing, we epoched the data prior to feature computation with *hctsa*. This step required the selection of an epoch length appropriate to both the data and to the application of *hctsa* to the data. In particular, the *hctsa* feature set requires regularly sampled time-series data without any missing data points (such as the discontinuities created by removing extreme artifacts during EEG pre-processing artifact rejection steps). Previous research has successfully used 30s epochs of EEG data with *hctsa* to characterise sleep stages (Decat et al., 2022). To ensure that as many participants as possible could be included without any data discontinuities, and to ensure data from all participants were consistent in length, 30s epochs (7500 samples) were selected for the current study, with the first artifact free 30s epoch from each participant’s recording used in the analysis. Removed electrodes were reconstructed using spherical spline interpolation (Perrin et al., 1989) to ensure that the same set of electrodes were present in all data. As many 30s epochs as possible were extracted from the cleaned data from each participant, starting from the start of the EEG file, with another epoch extracted starting from every subsequent 10s mark throughout the EEG data (so epochs were constructed with a 20s overlap). Participants that did not provide a single 30s epoch without any data discontinuities from their clean data were excluded from further analysis.

Data from each electrode from all epochs were baseline corrected to the average amplitude over the entire epoch and RELAX’s default epoch rejection settings were then applied, with the exception that an epoch was only rejected if the voltage shift within the epoch exceeded 120 microvolts (as visual inspection of the data showed that some participants had alpha oscillations that exceeded the typical 80 microvolt threshold). This first artifact-free 30s epoch was selected from each participant for inclusion in the analysis. Eight participants were excluded at this stage for not providing any artifact-free 30s epoch. Finally, the first epoch that remained after each of these steps was down-sampled to 250Hz to enable faster computation of the *hctsa* features, while still providing a sampling rate well above the 80Hz low-pass filter so that patterns can be detected even in the highest frequencies remaining in the data.

*hctsa* extracts features from a univariate time-series, whereas EEG data are multivariate (with data recorded from many electrodes simultaneously). To address this, we used PCA to reduce the multivariate dataset to components that explained significant variance within the EEG data, providing highly explanatory time-series from these principal components (PCs) (Bailey et al., 2023). To prevent the potential for class imbalance in EEG amplitudes to bias the PCA components towards individuals with larger amplitude EEG signals, we *z*-transformed each participant’s 26 electrode x 7500 timepoint data based on all values in the 30 s electrode x timepoint matrix. This normalised the overall voltage amplitude of each participant’s EEG data prior to the PCA but preserved potential relationships between electrodes (for example, occipital electrodes generally produce larger amplitude alpha activity). Next, we concatenated all participant data together across the time axis into a single 26 electrode x 1,410,000 time-samples matrix, performed a PCA decomposition on this matrix, and extracted the top components, which explained >85% of variance in the data (5 components). Concatenating all data prior to the PCA ensured all participants had the same PCA weights applied to their data, which were obtained from performing the PCA on the overall dataset. Using this consistent set of PCA loadings across participants, five PC time courses were extracted from each participant’s epoch for *hctsa* feature extraction. This resulted in a 5 principal component x 7500 sample multivariate time-series for each participant, reflecting a 30s extract of their cleaned data.

### *hctsa* computations and normalisation

Version 1.07 of *hctsa*, which includes implementations of 7729 time-series features, was used with MATLAB (2022b) to extract features and then fit and evaluate the classification models. The *hctsa* feature set includes time-series analysis methods developed in a range of scientific fields, including, physics, seismology, and economics (Fulcher & Jones, 2017; Fulcher et al., 2013). Each feature was computed for each participant and PC to produce a single value for each feature (two examples of features included in *hctsa* are the autocorrelation at a 3-sample lag, and the power in the lowest fifth of sampled frequencies in a Fourier power spectrum). Any non-real values or errors that were returned (due to an algorithm being inappropriate for a given dataset) were excluded. The time-series of the 188 participants who remained in the dataset after the artifact cleaning steps provided output values for more than 80% of the features as a real and non-error value, so all of these 188 participants were included in the analysis. Features were excluded if they did not produce real and well-behaved outputs across all participants. For example, feature computation for PC3 resulted in the removal of 425 features (out of 7729), with 7304 features remaining for subsequent analysis (represented as a 188 x 7304 participant x feature matrix).

Since the scale of each feature was specific to the time-series analysis performed, we normalised the *hctsa* feature data by performing a *z*-score transform across all participants within each feature separately, to enable more straightforward comparison of features measured on different scales and with different distributions. After normalisation, we used a linear support-vector machine (SVM) learning algorithm to classify ‘responder’/‘non-responder’ categories from the time-series features obtained from each PC component separately (represented as a 188 participant x 7304 feature matrix) (Fulcher & Jones, 2017; Fulcher et al., 2013). To account for class imbalance (with 66.5% responders), we used inverse probability class reweighting for the SVM training, and used leave-one-out cross-validation to obtain a balanced accuracy score to evaluate performance (Fulcher & Jones, 2017). To assess the potential clinical utility of our findings, we also report the normalised positive predictive value, which indicates the potential increase in response rates if treatment is provided only to patients who the algorithm predicts will respond (normalised by the response rate of the sample) (Krepel et al., 2019). To assess the statistical significance of our prediction accuracy, we used model-based permutation tests, with 1000 permutation-based null samples. This involved shuffling the ‘responder’/’non-responder’ labels prior to null model construction to create a distribution of null prediction accuracies for comparison to the balanced accuracy resulting from the model obtained from the real data (Henderson & Fulcher, 2022). This allowed us to derive a *p*-value as the proportion of null prediction accuracies that exceeded the real prediction accuracy (Fulcher & Jones, 2017). We controlled for multiple comparisons across the tests of the five tested PCs (from PCs 1-5) using the false discovery rate (FDR) (Benjamini & Hochberg, 1995), and report FDR corrected *p*-values (*p*_FDR_). Finally, we assessed the potential to differentiate responders and non-responders provided by individual features, to determine which features were providing any accurate predictions. This step is explained in the following section.

### Feature interpretation

As mentioned in the introduction, a considerable advantage of the *hctsa* method is that it highlights the types of interpretable time-series analysis methods that are informative for a given problem, in this case in distinguishing responders from non-responders from their EEG dynamics. While the SVM classification approach described above leverages thousands of time-series features simultaneously, for PC components with significant classification accuracy, we also wished to determine whether any time-series features were *individually* discriminative of responders and non-responders. To this end, we calculated the Mann–Whitney U test statistic (Mann & Whitney, 1947) for the differences between the responder and non-responder groups for each feature from any principal component that showed statistically significant classification accuracy. The Mann–Whitney U test is a non-parametric test that is robust to violations of normality, making no assumption about any specific distribution (McKnight & Najab, 2010). Due to the computation time involved in using the exact Mann–Whitney U test, we used the approximate test to begin with, which approximates the *p*-value from a Gaussian approximation of the distribution of the sum of ranks, an approach that is highly accurate for large samples (Cheung & Klotz, 1997). We then verified the accuracy of the approximate tests using the exact test (which computes the sum of ranks from the data without any approximation) for the 500 features that provided the strongest differentiation between the two groups (detected using the approximate test) to ensure the accuracy of the statistical tests of individual features with the strongest effect at separating the two groups. This allowed us to interpret which types of dynamical changes contained within a PCA component might underpin the classification accuracy. While the multivariate classifier using all *hctsa* features used normalized data from each feature (described above), in our individual feature analysis we analysed the raw (non-normalized) feature values to enable clearer interpretation of how each feature differed between the groups. We applied a FDR correction to control for multiple comparisons across all features using the method provided by Benjamini and Hochberg (1995). We note that this multiple comparison correction approach is likely to underestimate true feature significance, given the substantial correlations (and non-independence) between features (Fulcher et al., 2013).

Given this non-independence, we investigated the comparison of a reduced set of features by grouping highly correlated features using clustering, which reduced the number of required feature comparisons. We used *k*-medoids to cluster the features into an optimal number of clusters that reduced the substantial redundancy in the *hctsa* feature library and performed between-group comparisons of the principal variance within these clusters (Fulcher et al., 2013). To cluster the features, we used absolute Spearman correlation distances (Fulcher et al., 2013). The *k*-medoids algorithm initially assigned random data points as cluster centres, then iteratively assigned data points to their nearest cluster centre, re-computing cluster centres each time, and repeating this procedure until the algorithm converges on a stable solution (Park & Jun, 2009). This iterative process was performed across 100 iterations with 100 repeats to obtain the cluster that minimized the sum of distances of points to their nearest cluster centre. We implemented this approach for each possible number of *k*-medoid derived clusters from 2 to 100, then plotted the sum of the point-centroid distances for each cluster output from each of these potential number of clusters and selected the elbow in this plot as the number of clusters to test. The elbow suggested that 11 clusters was the optimal number of clusters to optimally represent the dataset (see Supplementary Materials Figure S5). After we obtained the list of features within each of these 11 clusters, we performed a PCA on the participant x feature matrix obtained by restricting the matrix to features identified as belonging to each cluster feature set separately, and extracted the component that explained the largest amount of the variance from within each cluster. This was used as the representative feature of each cluster, since taking the mean across the features within the cluster would not effectively represent the cluster due to anti-correlations between features within each cluster. We then compared the values from each of these 11 cluster-extracted components between the two groups using the Mann–Whitney U test and controlled for multiple comparisons across these 11 clusters using the method of Benjamini and Hochberg (1995).

### Sub-sample testing and independent validation dataset analysis

To determine whether our prediction accuracy was driven specifically by a particular sub-sample of the data, we performed a number of post-hoc tests where we examined the model’s balanced accuracy when restricted to only a sub-sample of the participants. Firstly, we tested balanced accuracy within only the participants who received left-hemisphere 10Hz rTMS treatment, and also within only the participants who received right-hemisphere 1Hz rTMS treatment. Secondly, we separately tested balanced accuracy within only the female and only the male participants.

After each of the previously described analyses were performed, we tested response prediction from using the PC and model that showed accurate prediction in the tests conducted on the full (*N* = 188) dataset applied to an independent validation dataset (*N* = 65, after pre-processing, N = 58, 30 responders). This independent validation dataset of participants was also from the TDBRAIN dataset, but only included participants who had been treated with 1Hz right hemisphere rTMS at a later date than our discovery dataset. Both the classification algorithm and the analyst (NWB) were blinded to the actual treatment outcome for this independent validation dataset. To obtain the model used for testing on the independent validation dataset, we trained the classification model on all participants in the discovery dataset. We applied the same RELAX data cleaning, epoching, baseline correction, *z*-transform and PCA weights from the full 188-participant discovery dataset to each of the participants in the validation dataset, so this data was transformed from raw data into processed data in exactly the same manner as the discovery dataset. We also applied the *hctsa* feature exclusions and *z*-score weightings from individual features in the discovery dataset to the independent validation dataset before using the model to predict the responder or non-responder identity of each participant from the independent validation dataset. Following these steps, individual responder and non-responder predictions were provided by the primary author (NWB) to HVD and MA who provided the balanced accuracy and positive/negative predictive values of the predictions. The primary author (NWB) then performed a permutation test to assess the statistical significance of the independent validation dataset balanced accuracy. This was performed by comparing the real balanced accuracy against a random (null) distribution of balanced accuracies. The random distribution was obtained by calculating the balanced accuracy from the prediction model applied to 1000 permutations of the labels from the independent validation dataset (with labels allocated in the same ratio as the real validation dataset).

## Results

### Leave-one-out cross-validation tests

PCs 1 to 5 explained 88.17% of the variance in the data (see Table 1 for the variance explained by each PC). PCs 1, 2, 4, and 5 did not show accurate prediction of treatment response (all balanced accuracies in the range of 45 to 55%, and all *p* and *p*_FDR_ > 0.10). However, the model that was trained on the PC3 time-series showed a balanced accuracy of 68.07% across the leave-one-out cross-validations (*p*_FDR_ = 0.005, see Figure 1 for a plot of the topography and power spectrum of PC3). The PC3 component contained a prominent alpha oscillation frequency peak, and a central-posterior maximum. The sensitivity was 85% and the specificity was 46%. The normalised-positive predictive value (PPV) was 114%, which reflects the increase in response rates if our classification algorithm had been used to select likely responders for treatment (Figure 2). This indicates a predicted increase of 14% in response rate compared to the observed response rate if participants had been allocated to receive rTMS based on the predictions made by our approach.

**Table 1.** The amount of variance explained by each of the first five principal components.

| Principal Component | Variance Explained |
| --- | --- |
| 1 | 50.07% |
| 2 | 16.61% |
| 3 | 13.45% |
| 4 | 4.36% |
| 5 | 3.68% |

**Figure 1.**
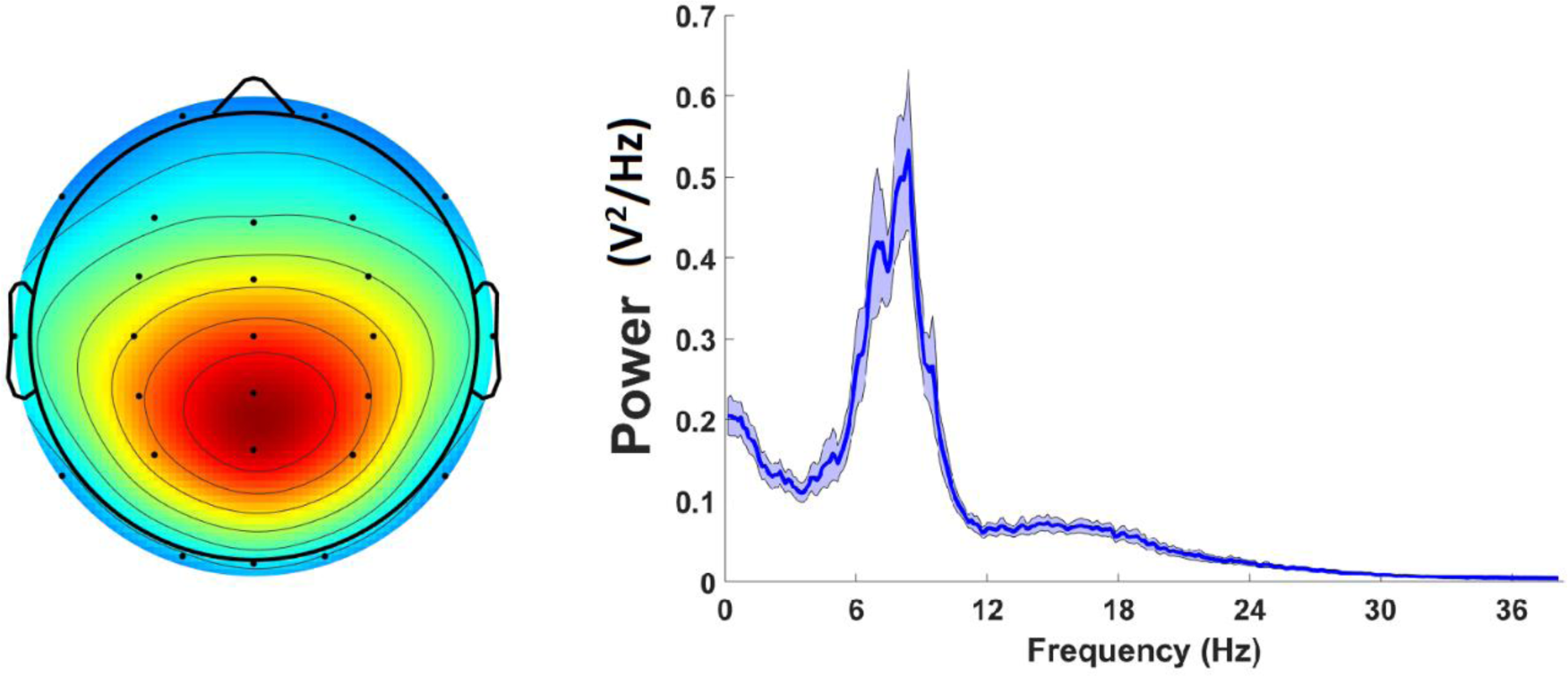
The topography and power spectrum for PC3. *Left*: The topographical map of PC3. *Right*: The power spectrum for PC3 averaged across all participants after a Welch transform within each participant individually, with the error bars reflecting 95% confidence intervals. The peak frequency after the power spectrum was averaged across all participants was 9.76Hz.

**Figure 2.**
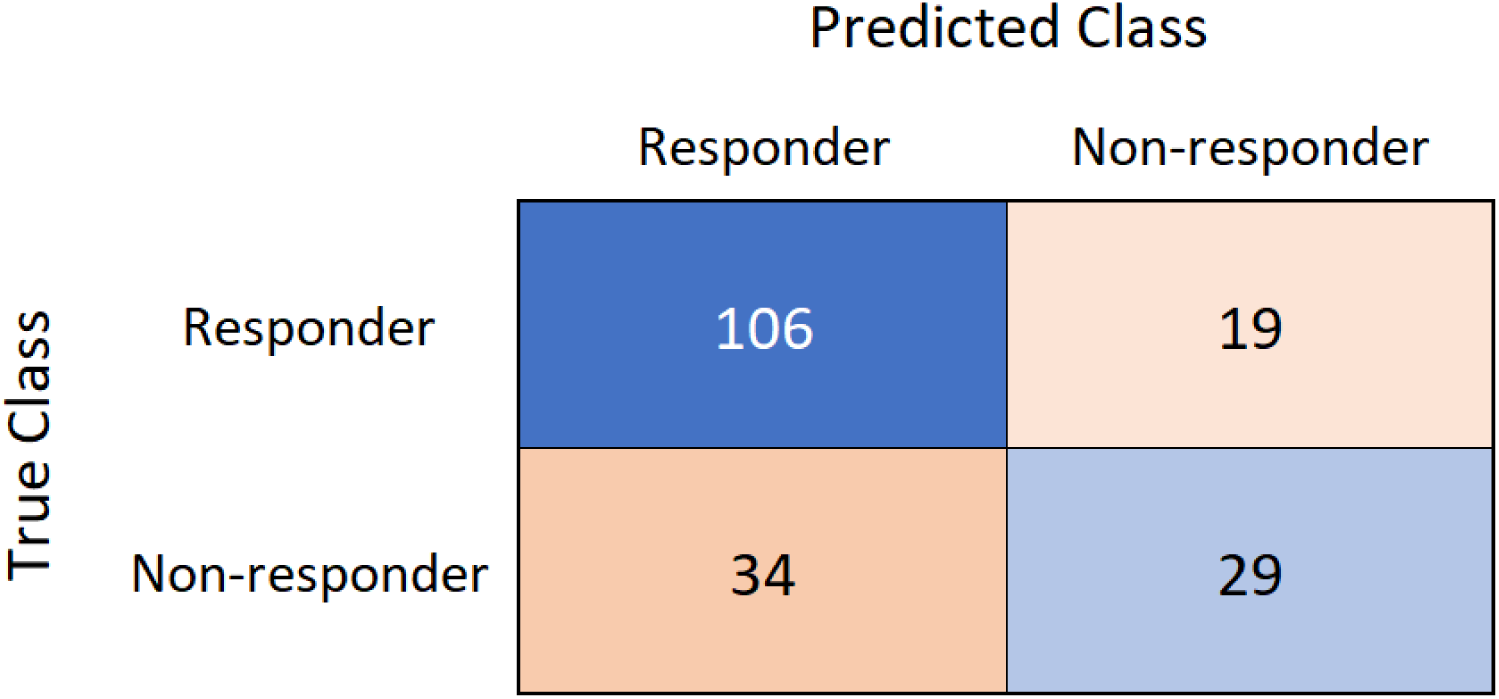
A confusion matrix of the predicted responder category from the leave-one-out cross-validation tests against true response or non-response categories in the discovery dataset.

### Which features of the time-series provide the response prediction?

Although PC3 showed above-chance classification accuracy, our analysis of individual features using the Mann–Whitney U test statistic showed that no single individual feature exceeded the FDR-corrected threshold for significance (all *p*_FDR_ > 0.05). This may be in part due to the substantial correlations between features, meaning that a much smaller number of independent tests were performed than our multiple comparison controls controlled for (Fulcher et al., 2013). In contrast, the results of our analysis of a reduced set of the primary variance from 11 clusters of features (representing 11 distinct concepts, with features within each of these 11 clusters providing highly related information, likely all representing a measure of a single underlying concept) indicated that two of these cluster extracted components were significant (*p*_FDR_ = 0.0379 and *p*_FDR_ = 0.0391). The first of these clusters contained features that indicated responders showed a negatively skewed distributional shape of their time-series values compared to non-responders, and the second cluster contained features indicating responders showed more high frequency power relative to low frequency power (see the supplementary materials for details). For the curious reader, we have provided a list and cluster plot of the top 50 features that showed the strongest effects for the difference between responders and non-responders in our supplementary materials, along with violin figures depicting a small number of representative features from these clusters. Similar to our cluster-based analysis of the full 7304 features restricted to 11 clusters of features, the cluster plot of the top 50 features showed two primary clusters. These clusters also represented features measuring the skew of the distributional shape and the high frequency power relative to low frequency power.

### Independent validation dataset

To assess the replicability of the prediction accuracy from the classification model described above we applied the overall model trained on all participants in the discovery dataset (*N* = 188) to the unseen independent validation dataset (*N* = 58, all treated with right-hemisphere 1Hz rTMS). The model predicted treatment response in the discovery participants with a balanced accuracy of 60% (permutation test *p* = 0.046). Given the predictions for the independent validation dataset, the normalised-PPV was 114%, a value that was identical to the value provided by our analysis of the discovery dataset. This normalised-PPV indicated a predicted increase of 14% in response rate compared to the observed response rate if participants had been allocated to receive rTMS based on the predictions made by our approach. This demonstrates the predictive accuracy of the model replicates in an independent dataset.

### Testing on sub-samples

In addition to the main tests above (conducted on the full sample) we also conducted some additional exploratory analyses to investigate whether our predictions were specific to one type of treatment, or might apply to one sex but not the other (as has been demonstrated in antidepressants by some previous research (Arns et al., 2016).

### rTMS protocol

Within the 71 participants who received left-sided 10Hz DLPFC treatment the time-series feature based classifier showed a mean balanced accuracy of 69.98% (*p* = 0.001). Within this left-sided 10Hz treatment population the normalised-PPV was 115%. The predictions for the 110 participants who received right-sided 1Hz DLPFC treatment yielded a mean balanced accuracy of 65.73% (*p* = 0.002). Within this right-sided 1Hz treatment population the normalised-PPV was 113%. This result demonstrates that the prediction accuracy from the overall discovery dataset classifier provided accurate predictions for both 1Hz right and 10Hz left-hemisphere treatments, rather than being specific to one treatment type.

### Sex differences

Within the 92 female participants the time-series feature based classifier showed a mean balanced accuracy of 71.04% (*p* < 0.001). Within the female sub-sample the normalised-PPV was 116%. The predictions for the 96 male participants showed a mean balanced accuracy of 65.33% (*p* = 0.006). Within the male sub-sample the normalised-PPV was 110%. This result demonstrates that the prediction accuracy from the overall discovery dataset classifier provided accurate predictions for both sexes, rather than being specific to one sex.

## Discussion

Our results indicate that a simple time-series feature-based classifier can predict rTMS treatment for depression from just 30s of baseline EEG data, using a posterior-central PC that was characterised predominantly by alpha activity and explained 13% of the variance in the data. This prediction ability was generalizable, remaining statistically significantly in an out-of-sample dataset (114% normalised-PPV, 60% balanced accuracy, *p* = 0.046), demonstrating modest potential of the approach for future clinical utility. Furthermore, our results showed this prediction accuracy was present for both 10Hz left-hemisphere rTMS treatment, and 1Hz right hemisphere rTMS treatment, and could be applied to both sexes.

The classification accuracy reported here is lower than other studies using baseline EEG to predict response to rTMS for depression. However, most previous research has not used an independent validation dataset to assess whether the accuracy obtained from the discovery dataset generalised to an independent dataset. Without this crucial step, we cannot determine the clinical utility of results – if the accuracy does not replicate out of sample, it will not provide accurate prospective prediction in the clinic. It is worth noting that our accuracy is within a similar range to previous research that has used an independent validation dataset test set to assess prediction accuracy from psychological symptoms and personality factors (Krepel et al., 2020). Furthermore, due to the comprehensive nature of *hctsa*, a vast range of potentially predictive single time-series EEG measures (including measures that were similar to those used in previous research) were assessed in the current study, which used a large dataset as well as an independent validation dataset test. Although our prediction accuracy was modest, we note that it was obtained from only 30 seconds of EEG data recorded from only 26 electrodes. Longer data periods with more electrodes may provide improved prediction accuracy. Interestingly, and in contrast to the majority of previous studies examining prediction of response to depression treatment research (Watts et al., 2022), our results showed better prediction of response to rTMS treatment for depression (sensitivity) than non-response (specificity). This might be clinically useful in providing a “go” signal to recommend rTMS treatment, providing a potential benefit over the typical ‘one-size-fits-all’ treatment recommendation approach, even in the context of modest prediction accuracy (Arns et al., 2022). The model reported in the current study might also be useful in combination with other high specificity models as an “ensemble” approach.

In addition to the response prediction accuracy, several points are worth noting. Firstly, our sub-sample analysis showed that responses to 10Hz left DLPFC treatment and 1Hz right DLPFC treatment could both be predicted accurately (balanced accuracy was 70% and normalised-PPV was 115% for 10Hz left DLPFC treatment compared to a balanced accuracy of 66% and normalised-PPV of 113% for 1Hz right DLPFC treatment). This is valuable, as it suggests our model works as a predictor of response to both commonly applied rTMS treatment approaches. However, further research could also explore the possibility of discriminant predictive measures. While our model was only trained on the dataset that included patients treated with rTMS applied to either left or right hemisphere, it may be that explorations of models and individual features that predict the right and left treatment responses separately could provide indications that patients will be more likely to respond to treatment to one hemisphere than the other. Patients could then be stratified to either 1Hz or 10Hz treatment based on which of the two sets of predictive measures provides the highest prediction of response (Arns et al., 2022; Garnaat et al., 2019). Since different treatments are likely to work via different mechanisms, measures that provide successful prediction for one treatment may predict non-response to another treatment. In this ideal situation, patients could be individually stratified based on their biotype to the treatment that best suited their biotype (Salomons et al., 2014). This stratification based on opposing patterns of response prediction has been demonstrated for attention deficit hyperactivity disorder, where low individualised alpha peak frequency predicted response to neurofeedback and high individualised alpha peak frequency predicted response to methylphenidate (Voetterl et al., 2022).

Additionally, with specific regards to the 10Hz left hemisphere treatment prediction accuracy, it is interesting to note that the principal component that provided the prediction accuracy was largely comprised of alpha activity, with an oscillatory frequency peak near to 10Hz. Previous research has indicated that the proximity of a patient’s peak alpha frequency to the 10Hz stimulation frequency relates to the response to left hemisphere 10Hz rTMS treatment (Roelofs et al., 2021), a finding that has been replicated (Voetterl et al., In Press). It may be that the relationships between the measures reported in the current study and treatment response are mediated by a similar underlying mechanism to the relationship between a patient’s peak alpha frequency and treatment response. Additionally, the results indicate that accurate response prediction was possible both within the female and male subgroups (although with slightly lower accuracy for males, with a normalised-PPV was 116% for females and 110% for males), suggesting generalisability to both sexes. It is not clear at this stage why prediction might be more accurate within females. However, we note that other research has also found higher predictive accuracy from EEG to antidepressant treatment within female participants (Arns et al., 2016).

In addition to our overall prediction accuracy, our analysis of the individual features suggests three conceptual points. First, when all individual features were compared between the groups and conservative multiple comparison controls were applied, none of the single features were able to provide statistical significance at distinguishing responders from non-responders. Inspection of the distributions for the highest performing individual features provided an explanation for this – the distributions of the two groups overlap, such that only a small minority of participants from each group fall outside of the distribution of the other group. As such, we could not extract statistically significant differentiation of responders and non-responders from individual features acting in isolation. Given the likely complex and subtle relationship between neural activity and response to rTMS treatment, this is perhaps unsurprising – it is unlikely that responders and non-responders differ in any specific quality such that they could be categorically separated without error, even if multimodal measurement techniques were to fully characterize each individual. This subtle relationship between neural activity and response to rTMS treatment may also explain why PC3, which only explained 13% of the variance in the EEG signal, provided prediction accuracy, while PC1 and 2 did not (despite containing much more of the variance). This suggests that the more dominant neural activity (reflected in PC1 and PC2) does not differ between responders and non-responders.

Second, the results indicate that the machine learning model could distinguish responders from non-responders with reasonable accuracy, even though no single feature could differentiate the two groups. This demonstrates the advantage of multivariate approaches that are trained on a large number of features such as *hctsa* or deep learning for accurate predictions over approaches trained on only a small number of features. To obtained clinically useful predictive accuracy, it seems important that many features of the data are used, with different weights provided to each feature based on the ability of the feature to distinguish the two groups. Predictions can be made for each individual based on the combination of the individuals’ feature values and the weightings, with the use of many measures providing the subtlety and a potential robustness in prediction accuracy that is not available from just a single feature.

Third, although no individual feature passed our conservative multiple comparison control, a restricted analysis of 11 clusters of highly correlated features (computed from these data using *k*-medoids clustering) indicated that two clusters were significant, passing our multiple comparison control threshold. These feature clusters suggested that within PC3, responders showed a pattern of more high frequency power relative to low frequency power, and a distribution of time-series values skewed towards negative deflections (with larger amplitude negative deflections) compared to non-responders. These types of features may be usefully explored by future research. In particular, the pattern whereby responders showed a distribution of time-series values that was skewed towards negative values is novel to TMS response prediction research, and is not typically examined in EEG research.

Finally, the modest prediction accuracy we detected is also consistent with our expectation that the true difference in EEG activity between responders and non-responders is small, and as such the modest accuracy reflects the plausibility of our results – highly accurate prediction of response would suggest a strong dependence between the baseline EEG measures (and only EEG measures) and the treatment response, in a manner that seems unlikely given the complexity and heterogeneity across individuals with depression in factors that might influence treatment response (Olbrich et al., 2015; Watts et al., 2022). While the *hctsa* model that showed significant classification accuracy may reflect a multivariate pattern that can provide modestly accurate prediction of response to rTMS, the relationship between this signal and treatment response may be variable, moderated by other (currently unknown) factors, and the relationship may not be present for all participants. Given the complexity and likely subtle relationship between potentially predictive baseline measures and response (Watts et al., 2022), even modest (above chance) prediction accuracy that translates to an out-of-sample dataset might still reflect a valuable contribution that could be developed with further research. It is worth noting here that prediction accuracy might be enhanced by using multimodal models that include behavioural or fMRI data in addition to the EEG data included in the current study.

### Limitations

The primary limitation of the current study is that the data was collected in the context of an uncontrolled open-label study. Previous research has demonstrated that fMRI-based connectivity can predict response to sham rTMS treatment, so it is possible our results are affected by non-specific effects (Wu et al., 2020). Participants were also medicated in many cases, which is likely to have affected the baseline brain activity sampled in our data. However, on the other hand, these aspects are also an advantage of the current study, as the design of the current study aligns with how rTMS treatment response prediction would be applied in the clinic (i.e., a naturalistic model), suggesting the translation of these findings into practice is plausible. The computation approach required to obtain the predictions in the current study also makes the translation of our results into practice plausible, as both the RELAX EEG pre-processing pipeline (Bailey et al., 2022a; Bailey et al., 2022b) and *hctsa* are freely available software (Fulcher et al., 2013), and the EEG data can be processed and *hctsa* feature set computed within 3-5 minutes per participant. However, although the pre-processing and prediction algorithm could be fully automated, clinics would still require access to EEG equipment and staff trained in the collection of EEG data.

Additionally, the independent validation dataset was collected across more recent years than the discovery dataset. Interestingly, the response rate to rTMS treatment for depression was lower in this more recent data, perhaps as a result of reductions in the non-specific effects aspect of the response as rTMS becomes more well-known and perhaps less novel for patients. As such, the independent validation dataset prediction balanced accuracy may have been lower than if our discovery dataset and our independent validation dataset were obtained during the same time period, although we note that the normalised PPV for the discovery and independent dataset were identical, indicating that the potential to improve rTMS response rates by predicting which participants would respond successfully generalised to the independent dataset. The overall model also performed worse at predicting response to right hemisphere 1Hz rTMS in our leave-one-out cross-validation tests, and the independent validation dataset only contained patients treated with right hemisphere 1Hz rTMS treatment. Prediction accuracy in our independent validation dataset may have been higher if the independent validation dataset contained participants treated with left-hemisphere 10Hz rTMS.

An additional limitation is that the EEG data analysed were only recorded at a single timepoint (before the start of rTMS treatment), and only the first 30s of artifact free EEG were analysed, so we did not assess the consistency of the EEG features across multiple recordings on different days, nor whether prediction accuracy might be provided by changes in time-series features during the course of treatment. Previous research has suggested that the change in non-linear dynamics within the alpha band from the first minute to the second minute of resting-state EEG data might predict non-response to rTMS treatment, and change in EEG connectivity after the first rTMS treatment predicts response, so predictions based on change in *hctsa* features across time may be worth exploring (Arns et al., 2014; Corlier, Wilson, et al., 2019). However, despite the minimal EEG data included and single timepoint measured, our predictive accuracy was significantly better than chance, suggesting that the EEG data provided features that were coupled with response to treatment weeks later. This indicates that there is a practical consistency across time in the data – the *hctsa* features must provide at least some information that relates across time to later treatment response. Another limitation is that the patients in the TDBRAIN dataset received psychotherapy concurrently with rTMS treatment, so our results might predict response to the combination of these two treatments rather than response to rTMS treatment alone, (although since patients presented to the clinic for TMS treatment suggests that the majority of participants were non-responders to prior psychotherapy alone). Data were also collected using a single model of EEG system, so it is not clear that results would generalise to data collected using other EEG systems.

One final limitation is that response prediction was implemented for a binary treatment response classification only, and only implemented for the end of the treatment. We note that there is a lack of consensus in how response should be defined (Watts et al., 2022). Some research has also suggested that different groups of response trajectories are apparent, with both fast and slow responder groups, and as such more fine grain in response prediction may improve predictive accuracy (Kaster et al., 2019). Alternatively, the maintenance of response prediction until a later follow-up period would be more clinically valuable, providing more confidence for treatment recommendations that might lead to lasting improvement (Ge et al., 2020; Hopman et al., 2021).

### Future Research and Conclusions

We would like to note that within our parameter selection steps (which were aimed at optimising our approach on EEG data), our attempts to predict the sex of participants from EEG data were outperformed by deep learning methods (Van Putten et al., 2018). One potential reason for this could be that the deep learning approach incorporated all electrodes into its training steps, rather than reducing the data to a separate single time-series as is required by the *hctsa* approach. As such, the deep learning approach may have benefitted from the inclusion of relationships or interactions between electrodes in its model, whereas *hctsa* did not. In support of this suggestion, some research has shown that relative levels of oscillatory power between different electrodes, or measures of alpha asymmetry between the hemispheres is predictive of depression treatment response (Arns et al., 2016; van der Vinne et al., 2021; Watts et al., 2022). Additionally, research that has used measures of neural connectivity (across both EEG and fMRI) have shown predictive accuracy (Corlier, Wilson, et al., 2019; Klooster et al., 2020; Salomons et al., 2014), and increasing evidence suggests depression is a disorder involving dysregulated connectivity (Ge et al., 2020). Indeed, meta-analysis of fMRI research has suggested that rTMS response prediction from connectivity measures obtained both from within and to the default mode network is more accurate than for other treatments, with responders showing higher baseline connectivity (Long et al., 2020). Even though the PCs measured in the current study are distributed components, they do not provide a measure of the interactions between brain regions. If connectivity measures improve prediction accuracy, then future work examining the interaction between pairs of time-series from brain regions implicated in depression may lead to better performance than our approach of reducing the data to a single time-series and examining that time-series across a comprehensive set of features. We note that a highly comparative approach to comparing pairwise interactions in time-series was recently developed, as the *pyspi* toolbox, which has been applied to EEG data (Cliff et al., 2022).

Additionally, we applied the *hctsa* toolbox only to test response prediction from rTMS treatment. It would be useful to determine if the toolbox is useful in predicting antidepressant medication response, and whether the same features or a different feature set provides accurate prediction in that case. As mentioned earlier with reference to 1Hz vs 10Hz rTMS, the ideal situation would be to find mutually exclusive predictive features so participants could be stratified to the optimal treatment for their characteristics, based on their baseline profile (Arns et al., 2022). The *hctsa* could also usefully be applied to examining potential mechanisms of rTMS treatment response – research could test which features differentiate baseline and post rTMS treatment EEG data to provide a data driven indication of which features are altered by successful treatment.

Further, it may be useful to combine the EEG predictors with other measures that have shown predictive promise. These might include previous treatment resistance, duration of current episode, age, cognitive measures, self-reported psychological symptoms, depression severity measures, benzodiazepine use, heart rate, and genetics (Beuzon et al., 2017; Brakemeier et al., 2007; Garnaat et al., 2019; Holtzheimer III et al., 2004; Kaster et al., 2019; Krepel et al., 2020; Lacroix et al., 2021; Rostami et al., 2017; Toffanin et al., 2022; Voetterl et al., 2021). The proximity of a patient’s peak alpha frequency to the 10Hz stimulation frequency has also been demonstrated to relate to response to left hemisphere 10Hz rTMS treatment (Roelofs et al., 2021), a finding that has been replicated (Voetterl et al., In Press). Combining multiple predictors in this way might further enhance predictive accuracy. We note here that both early change in cognition and early change in depression provide predictive potential (independently of each other) and combining these measures with those reported in the current study might provide very high predictive accuracy (Feffer et al., 2018; Hoy et al., 2012; Mirman et al., 2022; Mondino et al., 2020; Toffanin et al., 2022). However, to ideally minimize patient and clinical burden, accurate response from baseline (rather than after 1 week of treatment) would be optimal. Another alternative that might enhance predictive accuracy could be to restrict participants included in the model to specific depression subtypes, to reduce the heterogeneity of the sample and provide more consistent neural data on which to train the machine learning algorithm (Widge et al., 2019). However, this depends on effective subtyping of depression, an outcome that may be feasible but has not yet been reliably achieved (Widge et al., 2022).

Finally, with regards to potential clinical application, the current results should be replicated in a sample collected from an independent research group. Two replication approaches would be important to test. Firstly, using an identical EEG system and recording parameters as were used in the current study, to directly test whether our results replicate. Secondly, using a different EEG system and similar recording parameters to test generalisability to other hardware. If the first approach were to provide a positive result but the later does not, it may be important to use the specific hardware implemented in the current study for successful response prediction. If both replications show positive results, our approach could be generalised to other EEG systems for broad clinical application. We note that the PCA weightings used in the current study could not be applied to alternative EEG montages, so the PCA computation step would need to be performed separately for alternative EEG montages. Future research may also consider whether prediction accuracy could be obtained using only a small number of electrodes to reduce the potential clinical burden of the EEG. Electrodes of focus could be extracted from the PC weightings provided in the current study. An independent replication attempt could be performed in a naturalistic sample, in clinics that already perform rTMS treatments, if EEG data is collected at baseline over several years to assess a large dataset similar to the data reported in the current study.

In conclusion, our results indicate that a comprehensive set of time-series feature obtained from a brief period of baseline eyes-closed resting-state EEG data provided modestly accurate predictions of responses to rTMS treatment for depression in both leave-one-out cross-validation and an independent validation dataset dataset. Predictions were statistically superior to chance for both left and right hemisphere rTMS treatments, and for both sexes. These results were in the context of a priori selected analysis parameters and no hyper-parameter tuning of the classification algorithm. We suggest multiple avenues for future research to test for improved prediction accuracy, which we hope might lead to useful treatment stratification in the clinic.

## Data Availability

The TDBRAIN database and code are available on the Brainclinics Foundation website at www.brainclinics.com/resources and on Synapse at www.synapse.org/TDBRAIN.

## Supplementary Materials

### Parameter testing

In order to determine the EEG pre-processing and *hctsa* parameters for optimal application to predicting response to rTMS treatment for depression, we first tested which parameters were best suited to classifying the sex of each participant from the baseline EEG recordings from the more complete subset of the TDBRAIN dataset containing participants who were over 18 years old, using the *catch22* subset of *hctsa* (Lubba et al., 2019). *catch22* is a 22 feature subset of the full *hctsa* feature set that contains a representative sample of the features contained in the full *hctsa* feature set, and is comprised of features that are both minimally redundant and provide a minimal reduction in classification performance across a range of applications compared to the full *hctsa* feature set (Lubba et al., 2019). This parameter-testing subset of the TDBRAIN dataset contained N = 968, with 476 female participants, after excluding participants who were under 18 and excluding a number of files for noisy data. This provided us with an indication of which parameter settings were best for enabling accurate classification of different groups in EEG datasets, giving us a rationale for the methodology used in our primary study. We provide a brief description of the different parameters tested and their outcomes below, with each processing step and parameter configured to the same as the methods described in the main manuscript, except for the parameter being tested.

Firstly, we assessed which principal component would provide accurate classification of sex. We found the first principal component provided accurate (above chance) classification of sex in our initial test, and that this classification was more accurate than PC2 and PC3, so we conducted the rest of our tests using PC1. Following this, we tested which EEG pre-processing pipeline provided better classification results. We tested RELAX (Bailey et al., 2022a; Bailey et al., 2022b) and the pre-processing approach provided as the default with the TDBRAIN dataset (van Dijk, van Wingen, et al., 2022), assessing prediction accuracy from the first three principal components. We found that the RELAX pipeline provided 60% (+/− 5.43%) balanced accuracy in 10-fold cross validation tests, while the default approach provided 55% (+/− 8.62%) balanced accuracy. We note that many *hctsa* features are sensitive to artifacts, and that RELAX aims to clean all artifacts from the continuous data (including eye movements, muscle activity and all other biological and non-biological artifacts) while the default TDBRAIN artifact cleaning focuses only on cleaning drift and eye movements from the continuous data (with other artifacts intended for rejection in the epoching step). As such, while the default TDBRAIN artifact cleaning approach is likely to be suitable for many applications, RELAX performed better in combination with *hctsa* so was applied in the current study.

Next, we used the RELAX pre-processing approach, and tested different epoch lengths: 20, 30 and 45 seconds. We found that 45 second epochs required excluding more participants, as the data from 73 participants did not have a 45 second epoch without any extreme artifact. The 45 second epochs also did not provide higher classification accuracy than the 30 second epochs, with a balanced accuracy of 59% (+/− 4.26%) compared to the 60% (+/− 5.43%) for the 30 second epochs and 58% (+/− 4.47%) for the 20 second epochs. As such, we selected to use the 30 second epochs for our primary comparisons of interest.

Next, we tested whether down sampling the data from 500Hz to 250Hz (to enable faster computation of the *hctsa* features) would adversely affect the classification accuracy. We found that down sampling the data provided essentially the same classification accuracy as the 500Hz sampling rate, with the 250Hz down sampled data providing classification accuracy of 59% (+/− 4.26%) and the 500Hz data providing classification accuracy of 59% (+/− 4.42%). It is also worth noting here that RELAX applied a low-pass filter to the data at 80Hz (since the vast majority of neural activity is contained below the 80Hz range), so down sampling to 250Hz focuses *hctsa* more appropriately on the neural dynamics that are likely to be detected. As such, we selected to down sample our data to 250Hz for our primary analyses.

Finally, we tested whether eyes-open resting-state recordings might provide higher classification accuracy for sex than eyes-closed resting-state recordings. We found that eyes-closed recordings provided higher accuracy (59% +/− 4.26%) compared to the eyes-open recordings (57% +/− 4.12%). We also found that more participants had to be excluded when using eyes-open recordings (an extra 43 participants from the sex prediction dataset). As such, we selected to use eyes-closed recordings in our primary analysis to maximise our sample size and potential prediction accuracy. Given these tests, the pre-processing parameters that enabled the highest prediction accuracy of sex in the parameter testing dataset were using RELAX (Bailey et al., 2022a; Bailey et al., 2022b) to pre-process the data, using 30s epochs, down sampling the data to 250Hz, and using eyes-closed EEG recordings.

### Individual feature analysis

As mentioned in the text of our main manuscript, our analysis of individual features using the Mann–Whitney U test statistic showed that no single individual feature exceeded the FDR-corrected threshold for significance (all *p*_FDR_ > 0.05). This may be partially explained by the fact that the assumption of independence of the tests intrinsic to multiple comparison control methods is violated given the substantial correlations between features, so the multiple comparison controls likely underestimated the number of significant features in the data (Fulcher et al., 2013). In addition to the reduction of the meaningful feature to 11 features based on clustering from the original 7304 features, we have provided a cluster plot of the 50 features that showed the strongest effects for the difference between responders and non-responders (Figure S1). We have also provided violin plots for representative and clearly interpretable exemplar features from these primary clusters to aid with interpretation. To determine which types of time-series properties these features measured, we organised them using linkage clustering based on absolute Spearman correlations (|ρ|) between the values provided for each individual feature, implementing a cluster threshold of |ρ| > 0.75, which yielded groups of features that showed similar behaviour within our dataset. The cluster plot in Figure S1 depicts two primary clusters of highly correlated features.

First, the distributional shape of the values was negatively skewed in responders, suggesting their negative voltage deflections within PC3 were larger compared to non-responders. A representative feature from within this cluster was DN_Moments_3, which was within the top 100 most discriminative features, and measured skewness after a *z*-score transform is applied to the time-series data. This feature indicated that the responder group showed a distribution of their time-series values that was more negatively skewed than the non-responder group. Another representative feature for this cluster is skewness_pearson, which was present in the top 50 most discriminative features, and provides a measure of the extent and direction of the skew of the data (with the responder group on average showing a more negative skew compared to the non-responder group). While skewness_pearson can be influenced by the centre location (overall mean) of the time-series data, interpretation of the skewness_pearson feature in the context of DN_Moments_3 indicates the higher negative skew in the responder group was the driver for the difference (rather than differences in the mean of the data). See Figures S2 and S3 for violin plots depicting the distribution of these features for each group.

Second, features that represented the computation of the Fourier power spectrum (using the periodogram with a Hamming window), followed by the measurement of normalized power in the higher frequencies (i.e., relative to total power) indicated that responders showed greater relative high-frequency power compared to non-responders. See Figure S4 for a representational feature from within this significant cluster when clustering was performed by *k*-medoids across all individual features.

While features measuring these concepts showed the largest effect sizes in comparisons between the two groups, none of these features passed our (stringent) multiple comparison control threshold. Therefore, while it is likely that the combination of these features was driving the successful multivariate classification, we cannot be confident that any individual feature differentiated the two groups. However, we note that when the full feature set was reduced to 11 independent clusters, and the meaningful variance from each of these clusters were compared between the groups, our results showed that two of these clusters were significant (*p*_FDR_ < 0.05), and that these two clusters contained features similar to those reported from our top 50 features (and therefore likely represented the same concepts).

**Figure S1.**
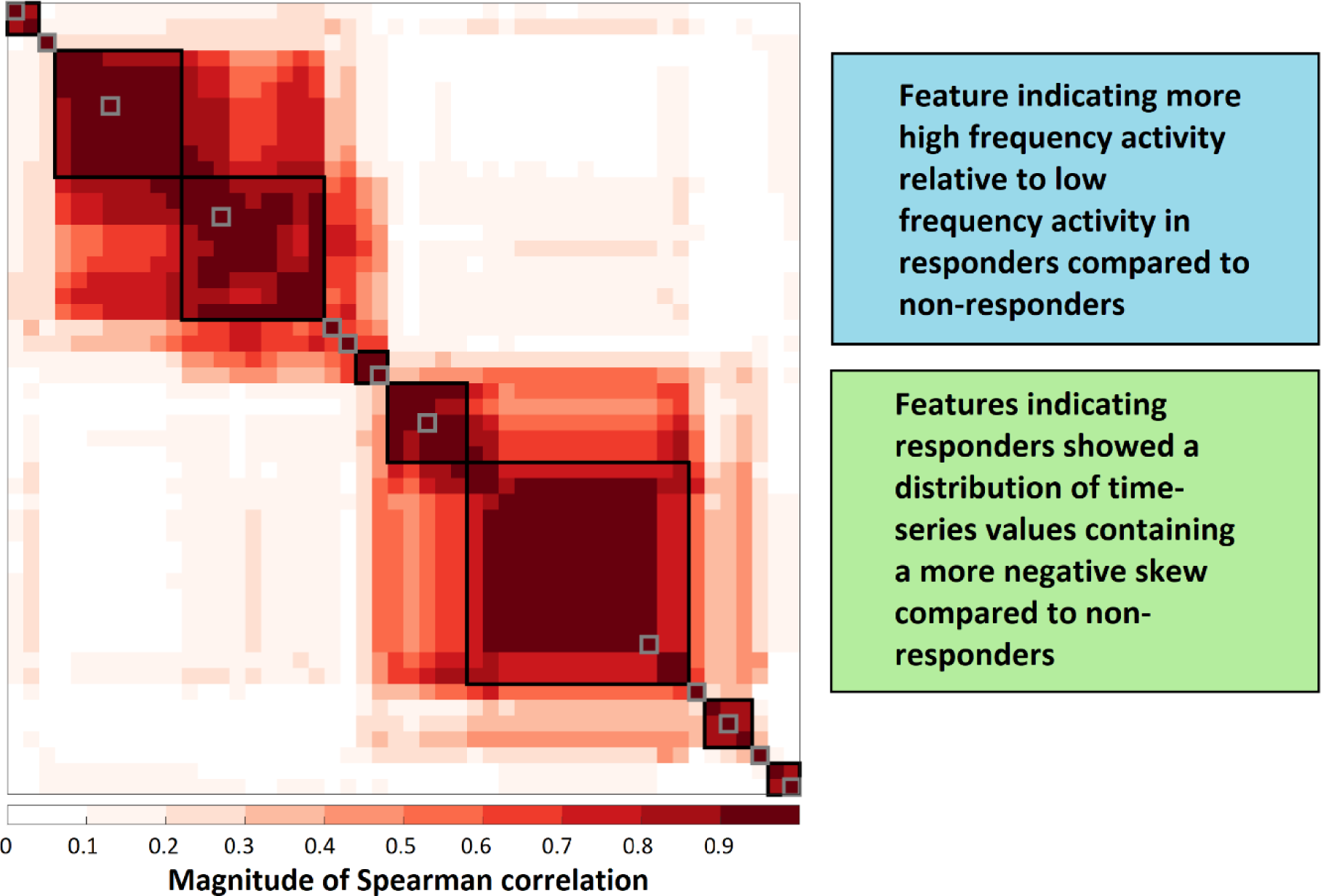
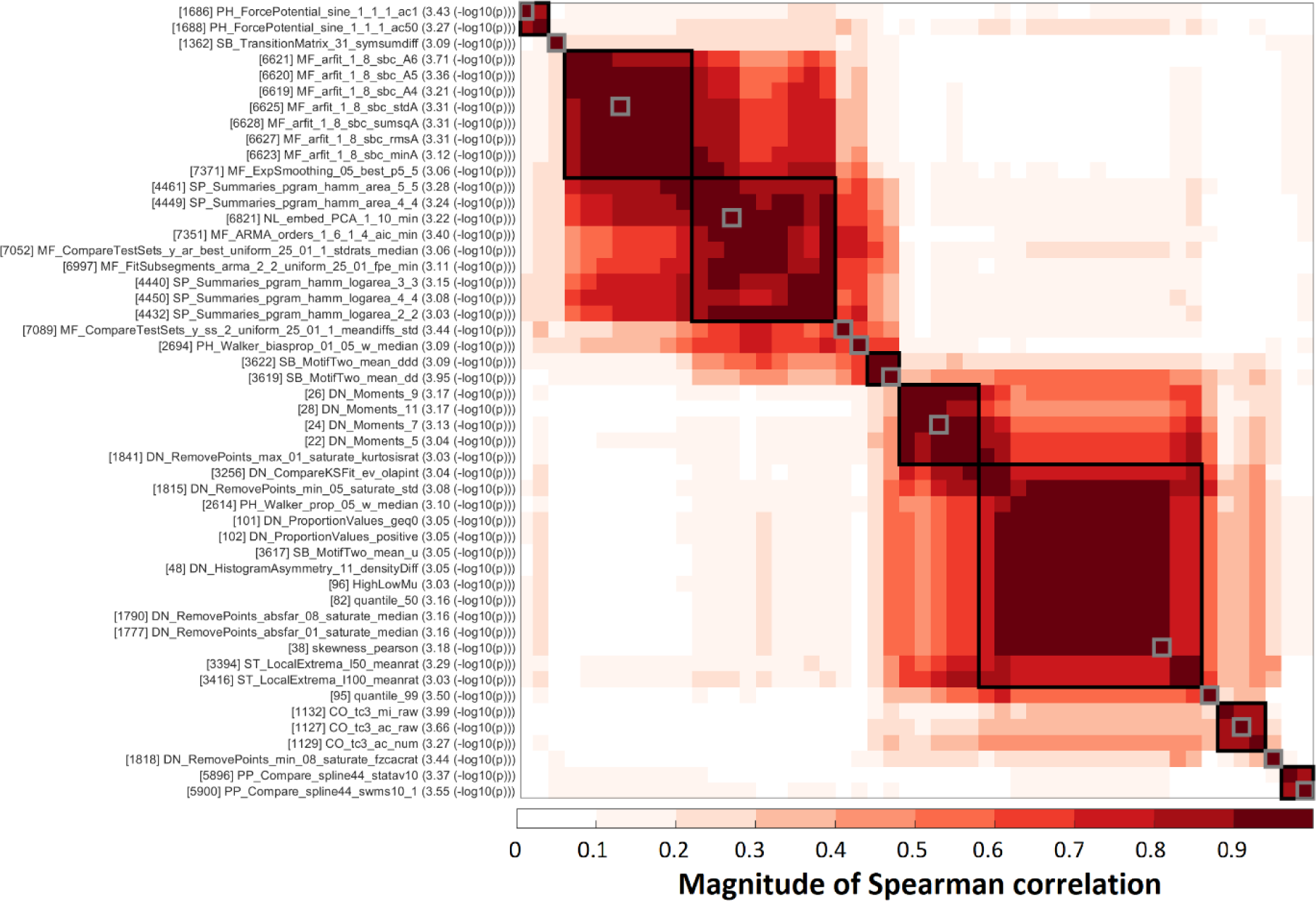
Top: A cluster plot of the top 50 features, with features clustered using absolute Spearman correlations and a threshold of |ρ| = 0.75 for forming clusters, and the primary clusters labelled by the concept they represent, enabling visualisation of how strongly the different clusters are related to each other. Bottom: The same cluster plot with individual feature labels provided.

**Figure S2.**
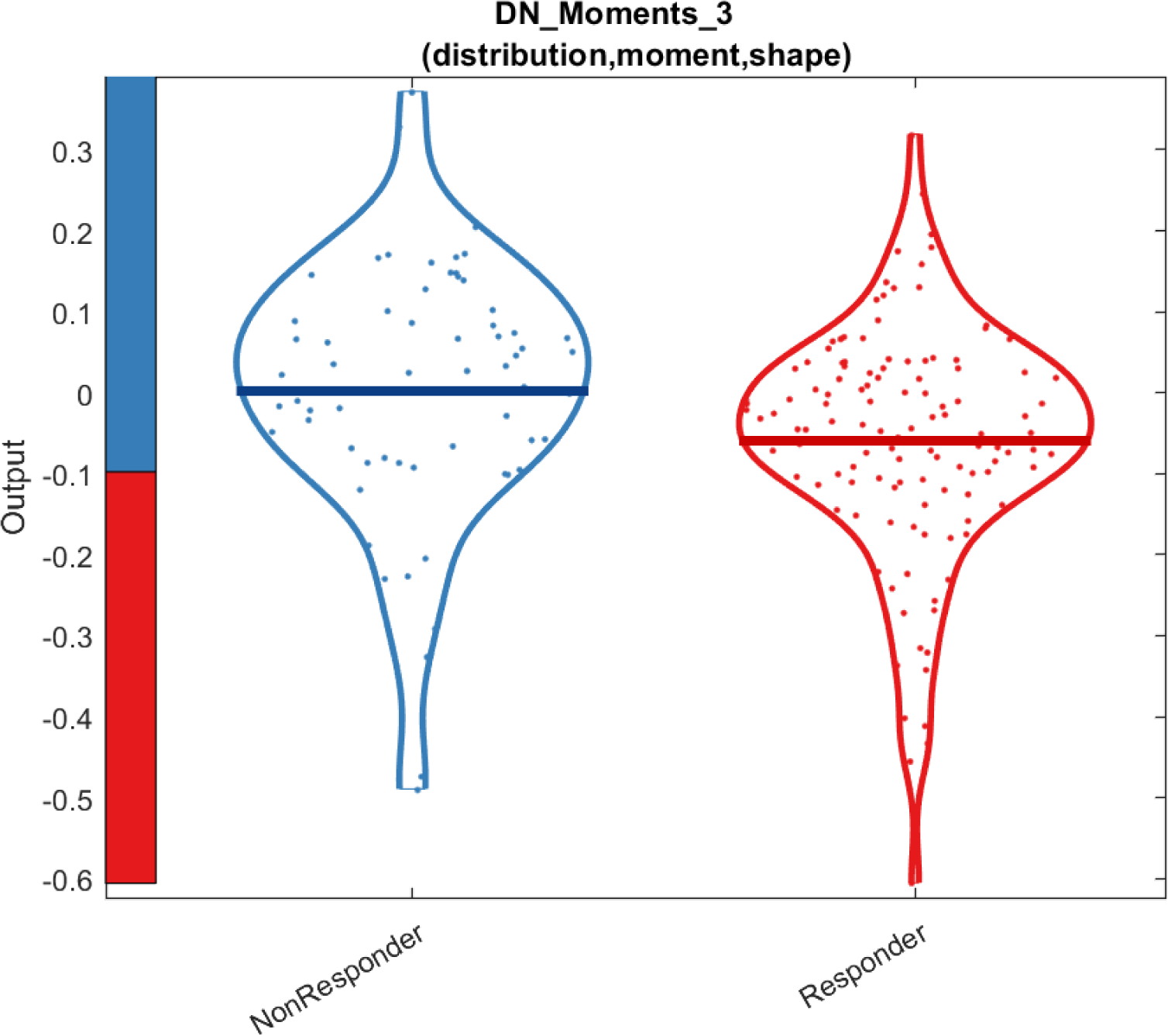
A violin plot for the DN_Moments_3 feature, which is a measure of skewness after a *z*-score transform is applied to the time-series data. This feature indicated responders showed a negative skew in their time-series data. While this feature did not pass our stringent FDR correction based multiple comparison controls, it was within a significant cluster after all features were separated into 11 representational clusters to reduce the redundancy in the *hctsa* feature list.

**Figure S3.**
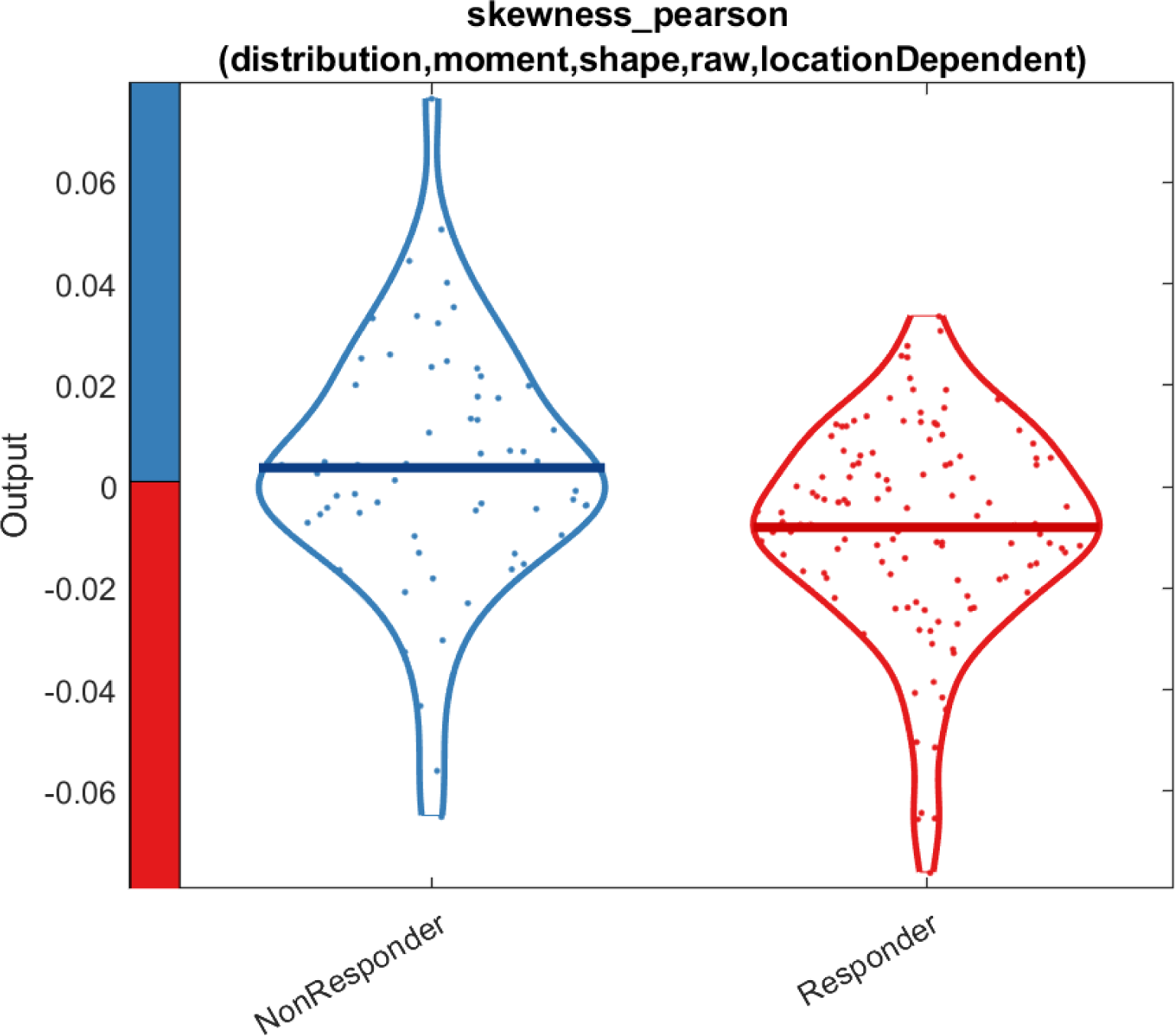
A violin plot for the measure of Pearson skewness. This feature indicated responders showed a negative skew in their time-series data. While this feature did not pass our stringent FDR correction based multiple comparison controls, it was within a significant cluster after all features were separated into 11 representational clusters to reduce the redundancy in the *hctsa* feature list.

**Figure S4.**
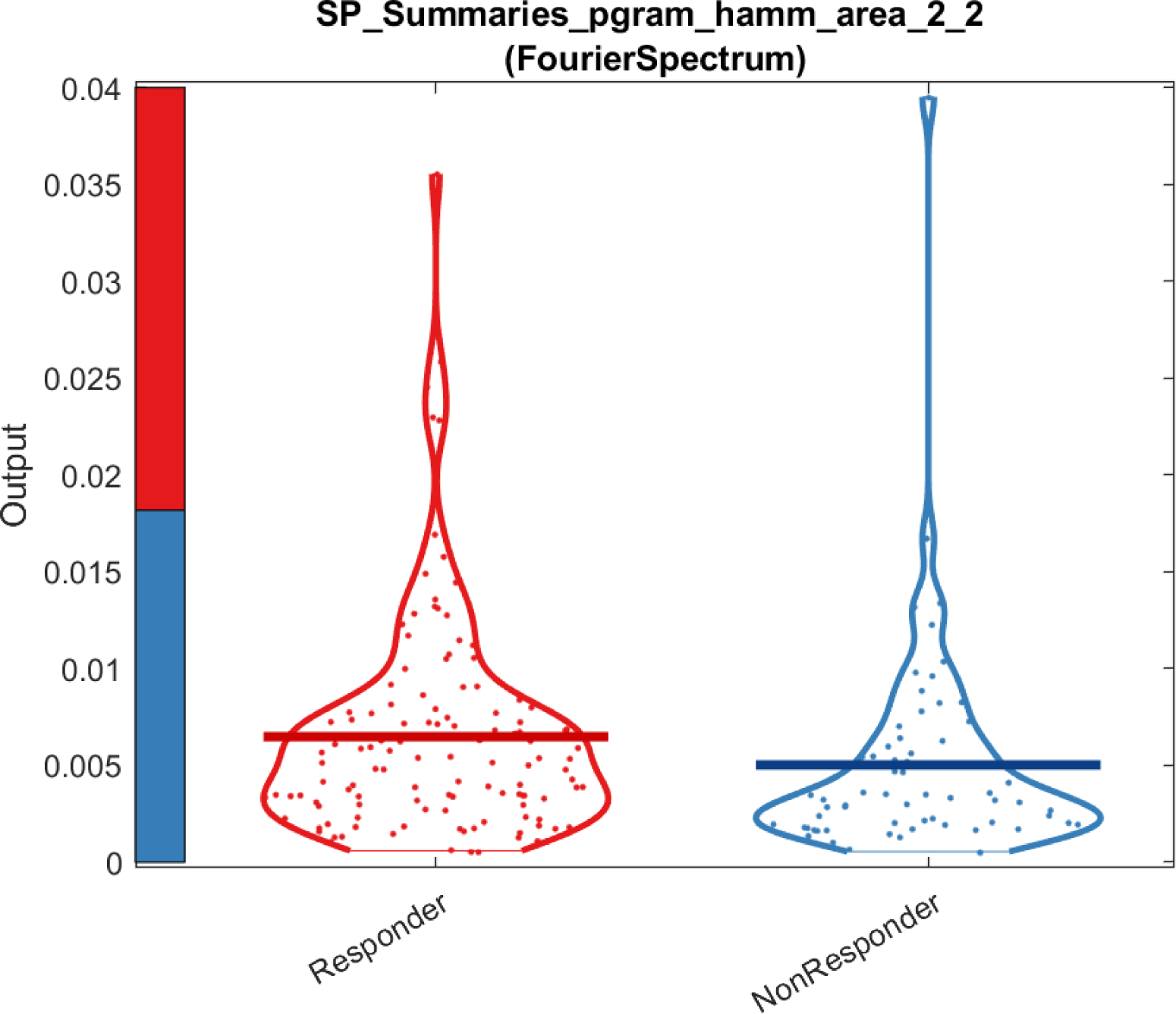
A violin plot for the power in the upper half of the frequencies measured using a periodogram with a Hamming window relative to the power in the lower half of the frequencies measured. This feature indicated responders showed a higher proportion of power in the upper half of frequencies measured than non-responders. While this feature did not pass our stringent FDR correction based multiple comparison controls, it was within a significant cluster after all features were separated into 11 representational clusters to reduce the redundancy in the *hctsa* feature list.

**Figure S5.**
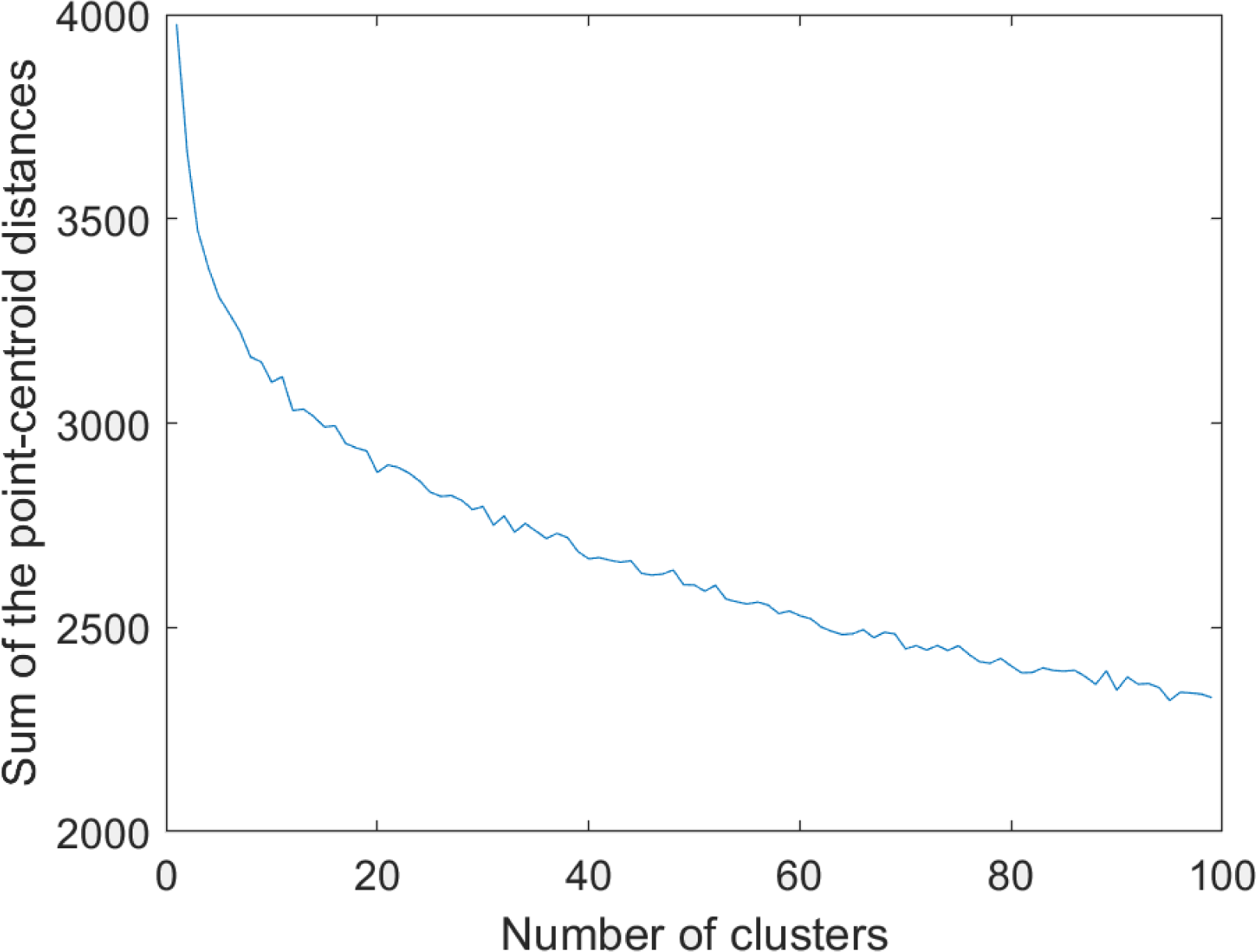
The sum of the point-centred distances between each feature and the centre point across each cluster for each number of clusters tested by k-medoids clustering. Note the elbow in the graph at approximately 11 clusters.

## Notes

**Funding Information:** PBF is supported by a National Health and Medical Research Council of Australia Investigator grant (1193596).

**Conflict of Interest:** In the last 3 years PBF has received equipment for research from Neurosoft, Nexstim and Brainsway Ltd. He has served on scientific advisory boards for Magstim and LivaNova and received speaker fees from Otsuka. He has also acted as a founder and board member for TMS Clinics Australia and Resonance Therapeutics. MA holds equity/stock in neurocare and Sama Therapeutics, serves as consultant to Synaeda, Sama therapeutics and Roche; Brainclinics Foundation received equipment support from neuroconn and Deymed. The other authors declare that they have no conflicts of interest.

### Competing Interest Statement

In the last 3 years PBF has received equipment for research from Neurosoft, Nexstim and Brainsway Ltd. He has served on scientific advisory boards for Magstim and LivaNova and received speaker fees from Otsuka. He has also acted as a founder and board member for TMS Clinics Australia and Resonance Therapeutics. MA holds equity/stock in neurocare and Sama Therapeutics, serves as consultant to Synaeda, Sama therapeutics and Roche; Brainclinics Foundation received equipment support from neuroconn and Deymed. The other authors declare that they have no conflicts of interest.

### Funding Statement

The research comprising the current study did not receive any direct funding. PBF is supported by a National Health and Medical Research Council of Australia Investigator grant (1193596).

### Author Declarations

We used a subset of the full publicly available TDBRAIN dataset that contained participants over 18 years of age (N = 968). All participants received the EEG as part of their routine care and provided informed consent for their data to be recorded and shared for the purposes of research.

## References

Arns, M., Bruder, G., Hegerl, U., Spooner, C., Palmer, D. M., Etkin, A., Fallahpour, K., Gatt, J. M., Hirshberg, L., & Gordon, E. (2016). EEG alpha asymmetry as a gender-specific predictor of outcome to acute treatment with different antidepressant medications in the randomized iSPOT-D study. Clinical neurophysiology, 127(1), 509–519.

Arns, M., Cerquera, A., Gutiérrez, R. M., Hasselman, F., & Freund, J. A. (2014). Non-linear EEG analyses predict non-response to rTMS treatment in major depressive disorder. Clinical neurophysiology, 125(7), 1392–1399.

Arns, M., van Dijk, H., Luykx, J. J., van Wingen, G., & Olbrich, S. (2022). Stratified psychiatry: Tomorrow’s precision psychiatry? European Neuropsychopharmacology, 55, 14–19.

Bailey, N., Biabani, M., Hill, A. T., Miljevic, A., Rogasch, N. C., McQueen, B., Murphy, O. W., & Fitzgerald, P. (2022a). Introducing RELAX (the Reduction of Electroencephalographic Artifacts): A fully automated pre-processing pipeline for cleaning EEG data-Part 1: Algorithm and Application to Oscillations. Clinical neurophysiology.

Bailey, N., Hill, A., Biabani, M., Murphy, O., Rogasch, N., McQueen, B., Miljevic, A., & Fitzgerald, P. (2022b). Introducing RELAX (the Reduction of Electroencephalographic Artifacts): A fully automated pre-processing pipeline for cleaning EEG data – Part 2: Application to Event-Related Potentials. Clinical neurophysiology. 10.1101/2022.03.08.483554

Bailey, N. W., Fulcher, B. D., Caldwell, B., Hill, A. T., Fitzgibbon, B., van Dijk, H., & Fitzgerald, P. B. (2023). Uncovering a stability signature of brain dynamics associated with meditation experience using massive time-series feature extraction. bioRxiv, 2023.2006. 2023.546355.

Bailey, N. W., Krepel, N., van Dijk, H., Leuchter, A. F., Vila-Rodriguez, F., Blumberger, D. M., Downar, J., Wilson, A., Daskalakis, Z. J., & Carpenter, L. L. (2021). Resting EEG theta connectivity and alpha power to predict repetitive transcranial magnetic stimulation response in depression: a non-replication from the ICON-DB consortium. Clinical neurophysiology, 132(2), 650–659.

Benjamini, Y., & Hochberg, Y. (1995). Controlling the false discovery rate: a practical and powerful approach to multiple testing. Journal of the Royal statistical society: series B (Methodological), 57(1), 289–300.

Berlim, M., Van den Eynde, F., Tovar-Perdomo, S., & Daskalakis, Z. (2014). Response, remission and drop-out rates following high-frequency repetitive transcranial magnetic stimulation (rTMS) for treating major depression: a systematic review and meta-analysis of randomized, double-blind and sham-controlled trials. Psychological medicine, 44(2), 225–239.

Beuzon, G., Timour, Q., & Saoud, M. (2017). Predictors of response to repetitive transcranial magnetic stimulation (rTMS) in the treatment of major depressive disorder. L’encephale, 43(1), 3–9.

Bigdely-Shamlo, N., Mullen, T., Kothe, C., Su, K.-M., & Robbins, K. A. (2015). The PREP pipeline: standardized preprocessing for large-scale EEG analysis. Frontiers in neuroinformatics, 9, 16.

Brakemeier, E.-L., Luborzewski, A., Danker-Hopfe, H., Kathmann, N., & Bajbouj, M. (2007). Positive predictors for antidepressive response to prefrontal repetitive transcranial magnetic stimulation (rTMS). Journal of psychiatric research, 41(5), 395–403.

Cao, X., Deng, C., Su, X., & Guo, Y. (2018). Response and remission rates following high-frequency vs. low-frequency repetitive transcranial magnetic stimulation (rTMS) over right DLPFC for treating major depressive disorder (MDD): a meta-analysis of randomized, double-blind trials. Frontiers in psychiatry, 9, 413.

Cash, R. F., Zalesky, A., Thomson, R. H., Tian, Y., Cocchi, L., & Fitzgerald, P. B. (2019). Subgenual functional connectivity predicts antidepressant treatment response to transcranial magnetic stimulation: independent validation and evaluation of personalization. Biological psychiatry, 86(2), e5–e7.

Castellanos, N. P., & Makarov, V. A. (2006). Recovering EEG brain signals: Artifact suppression with wavelet enhanced independent component analysis. Journal of neuroscience methods, 158(2), 300–312.

Cheung, Y. K., & Klotz, J. H. (1997). The Mann Whitney Wilcoxon distribution using linked lists. Statistica Sinica, 805–813.

Cliff, O. M., Lizier, J. T., Tsuchiya, N., & Fulcher, B. D. (2022). Unifying pairwise interactions in complex dynamics. arXiv preprint arXiv:2201.11941.

Corlier, J., Carpenter, L. L., Wilson, A. C., Tirrell, E., Gobin, A. P., Kavanaugh, B., & Leuchter, A. F. (2019). The relationship between individual alpha peak frequency and clinical outcome with repetitive transcranial magnetic stimulation (rTMS) treatment of major depressive disorder (MDD). Brain stimulation, 12(6), 1572–1578.

Corlier, J., Wilson, A., Hunter, A. M., Vince-Cruz, N., Krantz, D., Levitt, J., Minzenberg, M. J., Ginder, N., Cook, I. A., & Leuchter, A. F. (2019). Changes in functional connectivity predict outcome of repetitive transcranial magnetic stimulation treatment of major depressive disorder. Cerebral Cortex, 29(12), 4958–4967.

de Aguiar Neto, F. S., & Rosa, J. L. G. (2019). Depression biomarkers using non-invasive EEG: A review. Neuroscience & Biobehavioral Reviews, 105, 83–93.

Decat, N., Walter, J., Koh, Z. H., Sribanditmongkol, P., Fulcher, B. D., Windt, J. M., Andrillon, T., & Tsuchiya, N. (2022). Beyond traditional visual sleep scoring: massive feature extraction and unsupervised clustering of sleep time series. Sleep Medicine, 98, 39–52.

Dinga, R., Schmaal, L., Penninx, B. W., van Tol, M. J., Veltman, D. J., van Velzen, L., Mennes, M., van der Wee, N. J., & Marquand, A. F. (2019). Evaluating the evidence for biotypes of depression: Methodological replication and extension of Drysdale et al. 2017. NeuroImage: Clinical, 22, 101796.

Drevets, W. C., Price, J. L., & Furey, M. L. (2008). Brain structural and functional abnormalities in mood disorders: implications for neurocircuitry models of depression. Brain structure and function, 213, 93–118.

Drysdale, A. T., Grosenick, L., Downar, J., Dunlop, K., Mansouri, F., Meng, Y., Fetcho, R. N., Zebley, B., Oathes, D. J., & Etkin, A. (2017). Resting-state connectivity biomarkers define neurophysiological subtypes of depression. Nature medicine, 23(1), 28–38.

Feffer, K., Lee, H. H., Mansouri, F., Giacobbe, P., Vila-Rodriguez, F., Kennedy, S. H., Daskalakis, Z. J., Blumberger, D. M., & Downar, J. (2018). Early symptom improvement at 10 sessions as a predictor of rTMS treatment outcome in major depression. Brain stimulation, 11(1), 181–189.

Fitzgerald, P. B., Hoy, K. E., Anderson, R. J., & Daskalakis, Z. J. (2016). A study of the pattern of response to rTMS treatment in depression. Depression and anxiety, 33(8), 746–753.

Fulcher, B. D., & Jones, N. S. (2017). hctsa: A computational framework for automated time-series phenotyping using massive feature extraction. Cell systems, 5(5), 527–531. e523.

Fulcher, B. D., Little, M. A., & Jones, N. S. (2013). Highly comparative time-series analysis: the empirical structure of time series and their methods. Journal of the Royal Society Interface, 10(83), 20130048.

Garnaat, S. L., Fukuda, A. M., Yuan, S., & Carpenter, L. L. (2019). Identification of clinical features and biomarkers that may inform a personalized approach to rTMS for depression. Personalized medicine in psychiatry, 17, 4–16.

Ge, R., Downar, J., Blumberger, D. M., Daskalakis, Z. J., & Vila-Rodriguez, F. (2020). Functional connectivity of the anterior cingulate cortex predicts treatment outcome for rTMS in treatment-resistant depression at 3-month follow-up. Brain stimulation, 13(1), 206–214.

Henderson, T., & Fulcher, B. D. (2022). Feature-Based Time-Series Analysis in R using the theft Package. arXiv preprint arXiv:2208.06146.

Holtzheimer III, P. E., Russo, J., Claypoole, K. H., Roy-Byrne, P., & Avery, D. H. (2004). Shorter duration of depressive episode may predict response to repetitive transcranial magnetic stimulation. Depression and anxiety, 19(1), 24–30.

Hopman, H., Chan, S., Chu, W., Lu, H., Tse, C.-Y., Chau, S., Lam, L., Mak, A., & Neggers, S. (2021). Personalized prediction of transcranial magnetic stimulation clinical response in patients with treatment-refractory depression using neuroimaging biomarkers and machine learning. Journal of affective disorders, 290, 261–271.

Hoy, K. E., Segrave, R. A., Daskalakis, Z. J., & Fitzgerald, P. B. (2012). Investigating the relationship between cognitive change and antidepressant response following rTMS: a large scale retrospective study. Brain stimulation, 5(4), 539–546.

Hyvarinen, A. (1999). Fast and robust fixed-point algorithms for independent component analysis. IEEE transactions on Neural Networks, 10(3), 626–634.

Kar, S. K. (2019). Predictors of response to repetitive transcranial magnetic stimulation in depression: a review of recent updates. Clinical Psychopharmacology and Neuroscience, 17(1), 25.

Kaster, T. S., Downar, J., Vila-Rodriguez, F., Thorpe, K. E., Feffer, K., Noda, Y., Giacobbe, P., Knyahnytska, Y., Kennedy, S. H., & Lam, R. W. (2019). Trajectories of response to dorsolateral prefrontal rTMS in major depression: a THREE-D study. American Journal of Psychiatry, 176(5), 367–375.

Klooster, D., Voetterl, H., Baeken, C., & Arns, M. (2023). Evaluating Robustness of Brain Stimulation Biomarkers for depression: A Systematic Review of MRI and EEG Studies. Biological psychiatry.

Klooster, D. C., Vos, I. N., Caeyenberghs, K., Leemans, A., David, S., Besseling, R. M., Aldenkamp, A. P., & Baeken, C. (2020). Indirect frontocingulate structural connectivity predicts clinical response to accelerated rTMS in major depressive disorder. Journal of Psychiatry and Neuroscience, 45(4), 243–252.

Krepel, N., Rush, A. J., Iseger, T. A., Sack, A. T., & Arns, M. (2020). Can psychological features predict antidepressant response to rTMS? A Discovery–Replication approach. Psychological medicine, 50(2), 264–272.

Krepel, N., Sack, A. T., Kenemans, J. L., Fitzgerald, P. B., Drinkenburg, W. H., & Arns, M. (2018). Non-replication of neurophysiological predictors of non-response to rTMS in depression and neurophysiological data-sharing proposal. Brain Stimulation: Basic, Translational, and Clinical Research in Neuromodulation, 11(3), 639–641.

Lacroix, A., Calvet, B., Laplace, B., Lannaud, M., Plansont, B., Guignandon, S., Balestrat, P., & Girard, M. (2021). Predictors of clinical response after rTMS treatment of patients suffering from drug-resistant depression. Translational psychiatry, 11(1), 1–7.

Long, Z., Du, L., Zhao, J., Wu, S., Zheng, Q., & Lei, X. (2020). Prediction on treatment improvement in depression with resting state connectivity: a coordinate-based meta-analysis. Journal of affective disorders, 276, 62–68.

Lubba, C. H., Sethi, S. S., Knaute, P., Schultz, S. R., Fulcher, B. D., & Jones, N. S. (2019). catch22: CAnonical Time-series CHaracteristics: Selected through highly comparative time-series analysis. Data Mining and Knowledge Discovery, 33(6), 1821–1852.

Mann, H. B., & Whitney, D. R. (1947). On a test of whether one of two random variables is stochastically larger than the other. The annals of mathematical statistics, 50–60.

McKnight, P. E., & Najab, J. (2010). Mann-Whitney U Test. The Corsini encyclopedia of psychology, 1–1.

Mignan, A., & Broccardo, M. (2019). One neuron versus deep learning in aftershock prediction. Nature 574: E1–E3. In.

Mirman, A. M., Corlier, J., Wilson, A. C., Tadayonnejad, R., Marder, K. G., Pleman, C. M., Krantz, D. E., Wilke, S. A., Levitt, J. G., & Ginder, N. D. (2022). Absence of early mood improvement as a robust predictor of rTMS nonresponse in major depressive disorder. Depression and anxiety, 39(2), 123–133.

Mondino, M., Szekely, D., Bubrovszky, M., Bulteau, S., Downar, J., Poulet, E., & Brunelin, J. (2020). Predicting treatment response to 1Hz rTMS using early self-rated clinical changes in major depression. Brain stimulation, 13(6), 1603–1605.

Olbrich, S., van Dinteren, R., & Arns, M. (2015). Personalized medicine: review and perspectives of promising baseline EEG biomarkers in major depressive disorder and attention deficit hyperactivity disorder. Neuropsychobiology, 72(3-4), 229–240.

Park, H.-S., & Jun, C.-H. (2009). A simple and fast algorithm for K-medoids clustering. Expert systems with applications, 36(2), 3336–3341.

Perrin, F., Pernier, J., Bertrand, O., & Echallier, J. F. (1989). Spherical splines for scalp potential and current density mapping. Electroencephalography and clinical neurophysiology, 72(2), 184–187.

Pion-Tonachini, L., Kreutz-Delgado, K., & Makeig, S. (2019). The ICLabel dataset of electroencephalographic (EEG) independent component (IC) features. Data in brief, 25, 104101.

Roelofs, C. L., Krepel, N., Corlier, J., Carpenter, L. L., Fitzgerald, P. B., Daskalakis, Z. J., Tendolkar, I., Wilson, A., Downar, J., & Bailey, N. W. (2021). Individual alpha frequency proximity associated with repetitive transcranial magnetic stimulation outcome: An independent replication study from the ICON-DB consortium. Clinical neurophysiology, 132(2), 643–649.

Rostami, R., Kazemi, R., Nitsche, M. A., Gholipour, F., & Salehinejad, M. (2017). Clinical and demographic predictors of response to rTMS treatment in unipolar and bipolar depressive disorders. Clinical neurophysiology, 128(10), 1961–1970.

Salomons, T. V., Dunlop, K., Kennedy, S. H., Flint, A., Geraci, J., Giacobbe, P., & Downar, J. (2014). Resting-state cortico-thalamic-striatal connectivity predicts response to dorsomedial prefrontal rTMS in major depressive disorder. Neuropsychopharmacology, 39(2), 488–498.

Sehatzadeh, S., Daskalakis, Z. J., Yap, B., Tu, H.-A., Palimaka, S., Bowen, J. M., & O’Reilly, D. J. (2019). Unilateral and bilateral repetitive transcranial magnetic stimulation for treatment-resistant depression: a meta-analysis of randomized controlled trials over 2 decades. Journal of Psychiatry and Neuroscience, 44(3), 151–163.

Somers, B., Francart, T., & Bertrand, A. (2018). A generic EEG artifact removal algorithm based on the multi-channel Wiener filter. Journal of neural engineering, 15(3), 036007.

Squarcina, L., Villa, F. M., Nobile, M., Grisan, E., & Brambilla, P. (2021). Deep learning for the prediction of treatment response in depression. Journal of affective disorders, 281, 618–622.

Toffanin, T., Folesani, F., Ferrara, M., Murri, M. B., Zerbinati, L., Caruso, R., Nanni, M. G., Koch, G., Fadiga, L., & Palagini, L. (2022). Cognitive functioning as predictor and marker of response to repetitive transcranial magnetic stimulation in depressive disorders: A systematic review. General Hospital Psychiatry.

Traut, N., Heuer, K., Lemaître, G., Beggiato, A., Germanaud, D., Elmaleh, M., Bethegnies, A., Bonnasse-Gahot, L., Cai, W., & Chambon, S. (2022). Insights from an autism imaging biomarker challenge: promises and threats to biomarker discovery. NeuroImage, 255, 119171.

Van Der Donckt, J., Van Der Donckt, J., Deprost, E., Rademaker, M., Vandewiele, G., & Van Hoecke, S. (2022). Do Not Sleep on Linear Models: Simple and Interpretable Techniques Outperform Deep Learning for Sleep Scoring. arXiv preprint arXiv:2207.07753.

van der Vinne, N., Vollebregt, M. A., Rush, A. J., Eebes, M., van Putten, M. J., & Arns, M. (2021). EEG biomarker informed prescription of antidepressants in MDD: a feasibility trial. European Neuropsychopharmacology, 44, 14–22.

van Dijk, H., Koppenberg, M., & Arns, M. (2022). Towards Robust, Reproducible, and Clinically Actionable EEG Biomarkers: Large Open Access EEG Database for Discovery and Out-of-sample Validation. Clinical EEG and Neuroscience.

van Dijk, H., van Wingen, G., Denys, D., Olbrich, S., van Ruth, R., & Arns, M. (2022). The two decades brainclinics research archive for insights in neurophysiology (TDBRAIN) database. Scientific data, 9(1), 1–10.

Van Putten, M. J., Olbrich, S., & Arns, M. (2018). Predicting sex from brain rhythms with deep learning. Scientific reports, 8(1), 3069.

Voetterl, H., Miron, J.-P., Mansouri, F., Fox, L., Hyde, M., Blumberger, D. M., Daskalakis, Z. J., Vila-Rodriguez, F., Sack, A. T., & Downar, J. (2021). Investigating EEG biomarkers of clinical response to low frequency rTMS in depression. Journal of Affective Disorders Reports, 6, 100250.

Voetterl, H., van Wingen, G., Michelini, G., Griffiths, K. R., Gordon, E., DeBeus, R., Korgaonkar, M. S., Loo, S. K., Palmer, D., & Breteler, R. (2022). Brainmarker-I Differentially Predicts Remission to Various Attention-Deficit/Hyperactivity Disorder Treatments: A Discovery, Transfer, and Blinded Validation Study. Biological Psychiatry: Cognitive Neuroscience and Neuroimaging.

Watts, D., Pulice, R. F., Reilly, J., Brunoni, A. R., Kapczinski, F., & Passos, I. C. (2022). Predicting treatment response using EEG in major depressive disorder: A machine-learning meta-analysis. Translational psychiatry, 12(1), 1–18.

Widge, A. S., Avery, D. H., & Zarkowski, P. (2013). Baseline and treatment-emergent EEG biomarkers of antidepressant medication response do not predict response to repetitive transcranial magnetic stimulation. Brain stimulation, 6(6), 929–931.

Widge, A. S., Bilge, M. T., Montana, R., Chang, W., Rodriguez, C. I., Deckersbach, T., Carpenter, L. L., Kalin, N. H., & Nemeroff, C. B. (2019). Electroencephalographic biomarkers for treatment response prediction in major depressive illness: a meta-analysis. American Journal of Psychiatry, 176(1), 44–56.

Widge, A. S., Ellard, K. K., Paulk, A. C., Basu, I., Yousefi, A., Zorowitz, S., Gilmour, A., Afzal, A., Deckersbach, T., & Cash, S. S. (2022). Treating Refractory Mental Illness With Closed-Loop Brain Stimulation: Progress Towards a Patient-Specific Transdiagnostic Approach. Focus, 20(1), 137–151.

Wu, G. R., Wang, X., & Baeken, C. (2020). Baseline functional connectivity may predict placebo responses to accelerated rTMS treatment in major depression. Human brain mapping, 41(3), 632–639.

